# Real-world evaluation of AI-driven COVID-19 triage for emergency admissions: External validation & operational assessment of lab-free and high-throughput screening solutions

**DOI:** 10.1101/2021.08.24.21262376

**Authors:** Andrew A. S. Soltan, Jenny Yang, Ravi Pattanshetty, Alex Novak, Yang Yang, Omid Rohanian, Sally Beer, Marina A. Soltan, David R. Thickett, Rory Fairhead, CURIAL Translational Collaborative, Tingting Zhu, David W. Eyre, David A. Clifton

**Affiliations:** John Radcliffe Hospital, Oxford University Hospitals NHS Foundation Trust; RDM Division of Cardiovascular Medicine, University of Oxford; Institute of Biomedical Engineering, Dept. Engineering Science, University of Oxford; University of Oxford Medical School; The Queen Elizabeth Hospital, University Hospitals Birmingham NHS Foundation Trust; Institute of Inflammation and Ageing, University of Birmingham; Big Data Institute, Nuffield Department of Population Health, University of Oxford; NIHR Health Protection Research Unit in Healthcare Associated Infections and Antimicrobial Resistance at University of Oxford in partnership with Public Health England

**Keywords:** SARS-CoV-2, COVID-19, Artificial Intelligence, Machine Learning, Screening test, Validation, Operational evaluation, Diagnosis, Electronic Health Records, Emergency Department

## Abstract

**Background:** Uncertainty in patients’ COVID-19 status contributes to treatment delays, nosocomial transmission, and operational pressures in hospitals. However, typical turnaround times for batch-processed laboratory PCR tests remain 12-24h. Although rapid antigen lateral flow testing (LFD) has been widely adopted in UK emergency care settings, sensitivity is limited. We recently demonstrated that AI-driven triage (CURIAL-1.0) allows high-throughput COVID-19 screening using clinical data routinely available within 1h of arrival to hospital. Here we aimed to determine operational and safety improvements over standard-care, performing external/prospective evaluation across four NHS trusts with updated algorithms optimised for generalisability and speed, and deploying a novel lab-free screening pathway in a UK emergency department.

**Methods:** We rationalised predictors in CURIAL-1.0 to optimise separately for generalisability and speed, developing CURIAL-Lab with vital signs and routine laboratory blood predictors (FBC, U&E, LFT, CRP) and CURIAL-Rapide with vital signs and FBC alone. Models were calibrated during training to 90% sensitivity and validated externally for unscheduled admissions to Portsmouth University Hospitals, University Hospitals Birmingham and Bedfordshire Hospitals NHS trusts, and prospectively during the second-wave of the UK COVID-19 epidemic at Oxford University Hospitals (OUH). Predictions were generated using first-performed blood tests and vital signs and compared against confirmatory viral nucleic acid testing. Next, we retrospectively evaluated a novel clinical pathway triaging patients to COVID-19-suspected clinical areas where either model prediction or LFD results were positive, comparing sensitivity and NPV with LFD results alone. Lastly, we deployed CURIAL-Rapide alongside an approved point-of-care FBC analyser (OLO; SightDiagnostics, Israel) to provide lab-free COVID-19 screening in the John Radcliffe Hospital’s Emergency Department (Oxford, UK), as trust-approved service improvement. Our primary improvement outcome was time-to-result availability; secondary outcomes were sensitivity, specificity, PPV, and NPV assessed against a PCR reference standard. We compared CURIAL-Rapide’s performance with clinician triage and LFD results within standard-care.

**Results:** 72,223 patients met eligibility criteria across external and prospective validation sites. Model performance was consistent across trusts (CURIAL-Lab: AUROCs range 0.858-0.881; CURIAL-Rapide 0.836-0.854), with highest sensitivity achieved at Portsmouth University Hospitals (CURIAL-Lab:84.1% [95% Wilson’s score CIs 82.5-85.7]; CURIAL-Rapide:83.5% [81.8 - 85.1]) at specificities of 71.3% (95% Wilson’s score CIs: 70.9 - 71.8) and 63.6% (63.1 - 64.1). For 3,207 patients receiving LFD-triage within routine care for OUH admissions between December 23, 2021 and March 6, 2021, a combined clinical pathway increased sensitivity from 56.9% for LFDs alone (95% CI 51.7-62.0) to 88.2% with CURIAL-Rapide (84.4-91.1; AUROC 0.919) and 85.6% with CURIAL-Lab (81.6-88.9; AUROC 0.925). 520 patients were prospectively enrolled for point-of-care FBC analysis between February 18, 2021 and May 10, 2021, of whom 436 received confirmatory PCR testing within routine care and 10 (2.3%) tested positive. Median time from patient arrival to availability of CURIAL-Rapide result was 45:00 min (32-64), 16 minutes (26.3%) sooner than LFD results (61:00 min, 37-99; log-rank p<0.0001), and 6:52 h (90.2%) sooner than PCR results (7:37 h, 6:05-15:39; p<0.0001). Sensitivity and specificity of CURIAL-Rapide were 87.5% (52.9-97.8) and 85.4% (81.3-88.7), therefore achieving high NPV (99.7%, 98.2-99.9). CURIAL-Rapide correctly excluded COVID-19 for 58.5% of negative patients who were triaged by a clinician to ‘COVID-19-suspected’ (amber) areas.

**Impact:** CURIAL-Lab & CURIAL-Rapide are generalisable, high-throughput screening tests for COVID-19, rapidly excluding the illness with higher NPV than LFDs. CURIAL-Rapide can be used in combination with near-patient FBC analysis for rapid, lab-free screening, and may reduce the number of COVID-19-negative patients triaged to enhanced precautions (‘amber’) clinical areas.

## Background

Reducing nosocomial transmission of SARS-CoV-2 has been identified as a priority in safeguarding patient and healthcare staff safety, particularly as individuals with existing medical conditions are at greatest risk of severe illness and death ^1–5^. However, as the early clinical course of infection is often characterised by weakly-specific symptoms, and can be asymptomatic, viral testing is necessary to identify cases and is mandated for all UK hospital admissions.^4^

The mainstay of testing is batch-processed laboratory polymerase chain assay (PCR), which is imperfectly sensitive and requires specialist equipment^6–8^. Turn-around times have shortened throughout the pandemic, typically to within 12-24h in hospitals in high- and middle-income countries, but the interim uncertainty around patients’ COVID-19 status may contribute to treatment delays and postpone transfer to wards, thereby contributing to nosocomial transmission and operational strain. Novel rapid testing solutions have been adopted, including point-of-care (POC) PCR, loop mediated isothermal amplification, and lateral flow antigen testing (LFD), despite limitations in throughput and sensitivity^9,10^. Where POC PCR is available, use is typically constrained to time-critical decisions, such as surrounding emergency or transplant surgery, due to supply^11,12^. Moreover, although LFD is lab-free and highly specific (>99.5%) ^13,14^, allowing for a role in community case-finding^15^, multiple reports show more limited sensitivity (∼40%-70%)^16–18^ leading up to the U.S. Food and Drug Administration issuing a Class 1 recall on June 10, 2021^19^. A recent study evaluating performance amongst unscheduled hospital admissions confirmed high specificity (99.6%), but relatively low sensitivity (62%)^10^.

We recently demonstrated that an artificial-intelligence (AI) screening test (CURIAL-1.0) can rapidly detect COVID-19 amongst patients being admitted to hospital, by recognising SARS-CoV-2-induced abnormalities in routinely collected data^20^. A strength of our approach is the use of readily available blood test, blood gas & physiological measurements which are typically collected within 1h of presentation to hospitals in high- and middle-income countries, without requiring patient exposure to ionising radiation^21,22^. Explainability analyses revealed that features most informative to predictions were components of the Full Blood Count (FBC) and vital signs (Basophil count, Eosinophil count and Oxygen requirements), offering promise for clinically-guided optimisation to reduce prediction time.

Whereas many studies have investigated AI applications for diagnosis and prognosis during the pandemic^23–26^, key reviews highlight sector-wide methodological and reporting concerns that threaten generalisability, questioning the suitability of many models to-date for clinical use^27–29^. Reviewing the contribution of AI to the COVID-19 response, a recent editorial highlighted the promise of CURIAL-1.0 amongst other solutions to support patients during the pandemic, discussing the importance of high-quality validation studies inclusive of diverse patient populations^30^. Moreover, additional work quantifying benefits in the real-world clinical setting would demonstrate the clinical added-value of such approaches.

Accordingly, in this study we investigate efficacy, generalisability and real-world operational benefits of AI-driven COVID-19 screening in emergency departments, using insight from explainability analyses to improve generalisability and exploit advances in near-patient diagnostics for lab-free COVID-19 screening^31^. First, we externally validate two models with rationalised predictors, optimised separately for throughput (CURIAL-Lab; vital signs & routine blood tests) and speed (CURIAL- Rapide; vital signs & FBC only), across three independent UK NHS hospital trusts and prospectively for the second wave of the UK COVID-19 epidemic at Oxford University Hospitals. Next, we propose and investigate a novel clinical triage pathway using our AI models to enhance sensitivity of rapid antigen test (LFD)-based triage for unscheduled admissions. Lastly, we deploy CURIAL-Rapide alongside an approved point-of-care FBC analyser (OLO; SightDiagnostics, Israel) to provide rapid lab-free COVID-19 screening in the John Radcliffe Hospital’s Emergency Department (Oxford, UK), evaluating real-world performance and operational characteristics at a time of falling COVID-19 community prevalence in the UK^32^.

## Methods

### Diagnostic models to identify patients presenting with COVID-19

We updated our previously described model, designed to identify patients presenting to hospital with COVID-19 using vital signs, blood gas and routine laboratory blood tests (CURIAL-1.0^20^), with additional training data. The model was trained previously using clinical data from patients presenting to emergency and acute medical services at Oxford University Hospitals (OUH) between Dec 1, 2017 and April 19, 2020; additional data on all COVID-19-positive patient presentations to June 30, 2020 were added (Appendix B)^20^. This was performed to encompass all COVID-19 cases presenting to OUH during the ‘first wave’ of the COVID-19 pandemic (Supplementary Figure S1). OUH consist of four teaching hospitals, serving a population of 600 000, and provides tertiary referral services to the surrounding region. Routine blood tests were full blood count (FBC), urea, creatinine and electrolytes (U&Es), liver function tests (LFTs), coagulation and C-reactive protein (CRP), due to their ubiquity within existing emergency care pathways and rapid results, typically within around 1 h.

Next, we eliminated predictors with lower relative feature importances to improve generalisability across hospitals. CURIAL-Lab uses a focussed subset of routinely performed blood tests (FBC, U&Es, liver function tests, CRP) and vital signs (Table 1), eliminating the coagulation panel and blood gas which are not universally performed and are less informative^20^. Separately, we optimised for result-time, developing a minimalist model (CURIAL-Rapide) considering only predictors that can be obtained by the patient bedside (FBC and vital signs). We selected the FBC due to recent approval of a point-of-care haematology analyser (OLO, SightDiagnostics, Israel) with a result-time of 10 minutes, and as explanability analyses showed FBC components were most informative^31^. Models were trained using the OUH first-wave dataset described above.

**Table 1:**
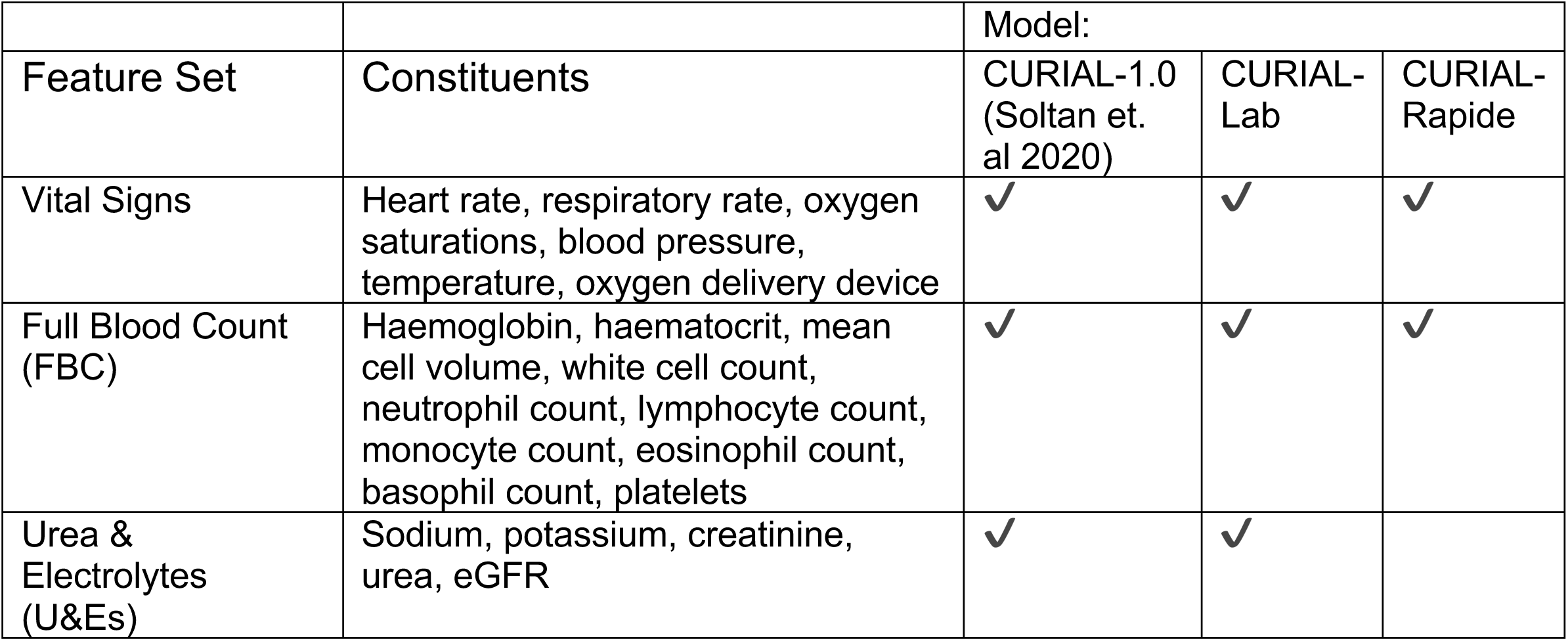
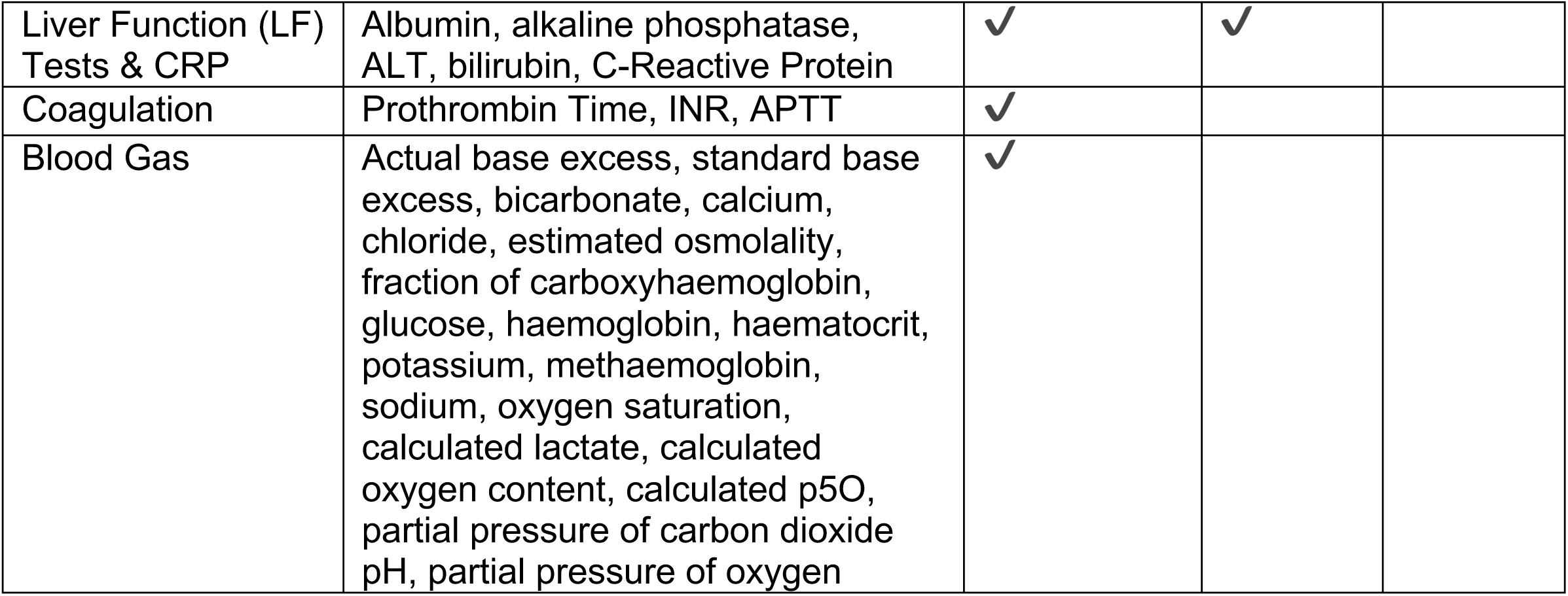
Clinical predictors considered in CURIAL-1.0, CURIAL-Lab & CURIAL-Rapide, showing successive elimination of less informative predictors to optimise for generalisbility and result-time. (ALT=alanine aminotransferase. APTT=activated partial thromboplastin time. CRP=C-reactive protein. eGFR=estimated glomerular filtration rate. INR=international normalised ratio. p50=pressure at which haemoglobin is 50% bound to oxygen. *c*=calculated)

| Feature Set | Constituents | Model: |  |  |
| --- | --- | --- | --- | --- |
|  |  | CURIAL-1.0<br>(Soltan et. al 2020) | CURIAL-Lab | CURIAL-Rapide |
| Vital Signs | Heart rate, respiratory rate, oxygen saturations, blood pressure, temperature, oxygen delivery device | ✓ | ✓ | ✓ |
| Full Blood Count (FBC) | Haemoglobin, haematocrit, mean cell volume, white cell count, neutrophil count, lymphocyte count, monocyte count, eosinophil count, basophil count, platelets | ✓ | ✓ | ✓ |
| Urea & Electrolytes (U&Es) | Sodium, potassium, creatinine, urea, eGFR | ✓ | ✓ |  |
| Liver Function (LF) Tests & CRP | Albumin, alkaline phosphatase, ALT, bilirubin, C-Reactive Protein | ✓ | ✓ |  |
| Coagulation | Prothrombin Time, INR, APTT | ✓ |  |  |
| Blood Gas | Actual base excess, standard base excess, bicarbonate, calcium, chloride, estimated osmolality, fraction of carboxyhaemoglobin, glucose, haemoglobin, haematocrit, potassium, methaemoglobin, sodium, oxygen saturation, calculated lactate, calculated oxygen content, calculated p5O, partial pressure of carbon dioxide pH, partial pressure of oxygen | ✓ |  |  |

### Evaluation for second wave of UK COVID-19 epidemic at OUH

We evaluated performance of CURIAL-1.0, CURIAL-Lab and CURIAL-Rapide using an independent prospective set of all patients presenting to emergency departments and acute medical services at OUH between October 01, 2020 and March 6, 2021, representing the second wave of the UK COVID-19 epidemic (Supplementary Figure 1). To assess performance characteristics at different calibrations, we performed the analysis with thresholds set during training to achieve sensitivities of 80% and 90%. Further, we performed a sensitivity analysis to assess susceptibility to imputation strategy, training the models using three independent imputation methods (median, mean and age-based mean) and examining variance in performance during evaluation. Mean sensitivity, specificity, area under receiver operator characteristic curve (AUROC) and predictive values are reported, alongside standard deviations across the three imputation methods. Confidence intervals for sensitivity, specificity and predictive values were computed using Wilson’s Method^33^, and for AUROC with DeLong’s method^34^.

### External validation at independent NHS trusts

We externally validated CURIAL-Rapide and CURIAL-Lab, calibrated during training to a sensitivity of 90%, across three independent UK National Health Service (NHS) hospital Trusts by comparing model predictions to confirmatory molecular testing (SARS-CoV-2 laboratory PCR, SAMBA-II and Panther). Participating hospital trusts were University Hospitals Birmingham NHS Foundation Trust (UHB), Bedfordshire Hospitals NHS Foundation Trust (BH), and Portsmouth University Hospitals NHS Trust (PUH), serving a total population of ∼3.5 million. We evaluated the models for all patients aged over 18 who had an unscheduled admission via emergency or acute medical pathways and received a blood draw on arrival during the specified date ranges. Screening against eligibility criteria, followed by anonymisation, were performed by the respective NHS trusts. Patients who dissented to EHR research, did not have confirmatory molecular testing for COVID-19, or had only an invalid confirmatory test result with no subsequent valid result, were excluded. For trusts where blood-gas results were available for electronic extraction, we also evaluated CURIAL-1.0. Evaluation periods and confirmatory testing method are listed in Appendix C.

### Comparison with Lateral Flow Devices

To investigate suitability of CURIAL-Rapide, CURIAL-Lab and CURIAL-1.0 as rapid screening tests for unscheduled admissions, we compared sensitivities and negative predictive values with LFD results from the Innova SARS-CoV-2 Antigen Rapid Qualitative Test which was used within the standard-of-care at OUH during the second wave of the UK COVID-19 epidemic (to March 6, 2021; Supplementary Figure S2). From December 23, 2020 patients admitted to OUH from acute and emergency care settings (Emergency Department, Ambulatory Medical Unit, Medical Assessment Unit) had LFDs performed routinely alongside PCR testing. Swabs of the nose and throat were collected for both tests by trained nursing or medical staff and LFDs were performed in the emergency/acute departments. Positive, negative and invalid LFD results were documented in the EHR. Swabs for PCR were transferred to the clinical laboratory in viral transport medium and tested by PCR (ThermoFisher TaqPath), forming the reference standard for evaluating model predictions and LFDs.

### A combined algorithm to enhance the sensitivity of LFD testing

Next, we investigated whether CURIAL-Rapide, CURIAL-Lab and CURIAL-1.0 could enhance the sensitivity of LFDs for identifying COVID-19 amongst patients being admitted to hospital. We proposed and retrospectively evaluated a novel clinical triage pathway (Figure 1) labelling patients as COVID-19-suspected where they had either a positive CURIAL result or a positive LFD result. Due to high specificity, in our pathway patients with positive LFDs can be streamed directly to a COVID-19-positive clinical area, meanwhile patients with a negative LFD but a positive CURIAL result would be managed in an enhanced-precautions area pending PCR adjudication. The pathway aimed to enhanced negative predictive value for patients receiving both negative LFD and CURIAL results, reduce the false-negative rate and therefore supporting safe and rapid triage directly to a ‘green’ zone.

**Figure 1:**
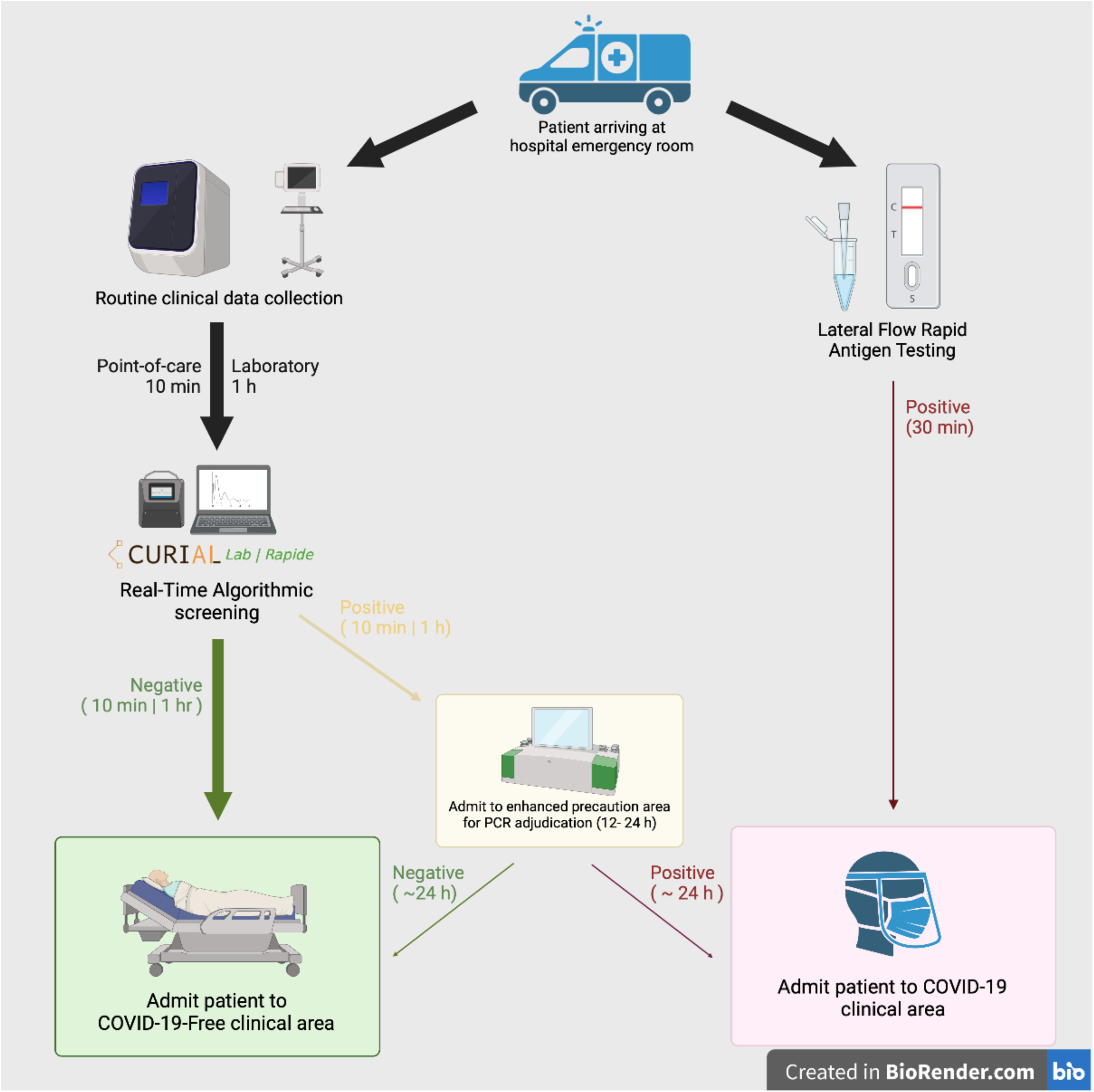
A rapid COVID-19 screening pathway for Emergency Department arrivals (a) routine blood tests and vital signs recordings are performed on patients’ arrival to the emergency department, either using rapid point-of-care haematology analysers (∼10 minutes; CURIAL-Rapide), or the existing laboratory analysis (∼1h; CURIAL-Lab). (b) Real-time algorithmic analysis of blood tests & vital signs allows early, high-confidence identification of negative patients for safe streaming to COVID-19-Free clinical areas. (c) Patients with positive screening test results are admitted to enhanced precautions (amber) areas, pending confirmatory PCR result. (d) Patients testing positive via Lateral Flow Test can be streamed directly to COVID-19 (‘red’) clinical areas. Arrow thickness represents patient flow.

We assessed performance of this novel pathway retrospectively for all unscheduled admissions to OUH where patients received LFD testing, from introduction on December 23, 2020 to March 6, 2021. Comparison between the performances of CURIAL-Lab, CURIAL-Rapide and CURIAL-1.0 alone are performed with integrated clinical pathway for each model using McNemar’s Chi-Square test.

The study protocol for model development, external and prospective validation was approved by the National Health Service (NHS) Health Research Authority (IRAS ID 281832) and sponsored by the University of Oxford.

### Prospective evaluation of CURIAL-Rapide in a lab-free clinical pathway

To prospectively assess operational and predictive performance of CURIAL-Rapide in a lab-free setting, we deployed two OLO rapid haematology analysers [SightDiagnostics, Tel Aviv] in the Emergency Department (ED) at the John Radcliffe Hospital, Oxford, as part of an OUH-approved service evaluation (Ulysses ID: 6907)^31^. We simultaneously aimed to improve routine clinical care by reducing the time for routine blood test results to become available in ED. The analysis plan and data requirements for the CURIAL-Rapide evaluation were determined prospectively and registered with the Trust service evaluation database.

We estimated a suitable review-point using Buderer’s standard formulae^35^. Predicting a sensitivity of CURIAL-Rapide of 80% (matching model calibration), specificity of 75%, and prevalence of COVID-19 at 15% amongst patients in ED, we estimated a minimum sample size of 410 enrolled patients to determine sensitivity and 85 patients to determine specificity (95% confidence, precision 10%^36^). We therefore planned to review model performance once 500 patients had been enrolled, to allow for missing or invalid confirmatory tests.

The service evaluation operated from February 18, 2021 to May 10, 2021 between 8am and 8pm for patients meeting the eligibility criteria. Patients eligible were aged over 18, attending the ED with an acute illness and streamed to a bedded clinical area, and had consented to receive full blood count analysis and vital signs as part of their emergency care plan. We selected patients allocated to bedded clinical areas as non-ambulatory patients typically have higher acuity of illness, therefore being more likely to benefit from faster blood test results and having a higher probability of admission. Patients were identified on arrival using the FirstNet system (Cerner Millennium, Cerner, UK).

Eligible patients were enrolled for additional near-patient, lab-free full blood count analysis using the OLO, which in conjunction with vital signs were used to generate CURIAL-Rapide predictions. OLO FBC results were uploaded immediately to the EHR, making results available to clinicians and supporting routine care. We excluded patients with an invalid OLO result and no subsequent successful result, thereby ensuring data completeness. Routine COVID-19 testing was performed in line with trust policies, with LFDs (Innova SARS-CoV-2 Antigen Rapid Qualitative Test) performed in the department and paired multiplex PCR on-premises in a dedicated laboratory (ThermoFisher TaqPath). Patients who did not have confirmatory testing for COVID-19 within routine care were excluded from performance evaluation.

We recorded patients’ arrival time to the hospital, measurement time of vital signs, and result times for LFD, PCR, OLO and laboratory FBC analysis. We also recorded the first-attending clinician’s triage impression of COVID-19 status, using the locally adopted Green/Amber/Blue categorization system (Green representing a patient whose illness has no features of COVID-19, amber representing an illness with features potentially consistent with COVID-19, and blue representing laboratory confirmed COVID-19 infection)^37,38^. Where COVID-19 triage category had not been documented by the first-assessing physician, adjudication was performed through clinical review of notes using rules-based determination. Patients having documentation of a new continuous cough, temperature ≥37.8°C, or loss or change in sense of smell or taste were adjudicated as an ‘amber’ (COVID-19-suspected) stream, matching UK Government guidance on definition of a possible COVID-19 case^8^. Patients with PCR-confirmed COVID-19 in the 10 days preceding attendance were adjudicated to the ‘Blue’ (COVID-19-confirmed) stream. Patients with no features of COVID-19 and no documented clinical suspicion were adjudicated to the ‘Green’ stream.

We generated CURIAL-Rapide predictions using OLO results and vital signs, comparing predictions against results of confirmatory PCR testing. CURIAL-Rapide predictions were not made available to the attending clinician so as not to influence the clinical triage category or decisions to proceed to confirmatory testing for patients being discharged. Availability time for CURIAL-Rapide was the later of OLO result time and vital signs recording time as both are required to generate a prediction. The time-to-result for PCR, lateral flow, and CURIAL-Rapide tests were calculated as the time from a patient’s first arrival in the ED to the time of a test result being available on the EHR.

We selected time-to-result as our primary outcome, recognising the role of rapid test results in reducing nosocomial transmission. Our secondary outcomes were sensitivity, specificity, PPV and NPV for CURIAL-Rapide & LFDs, and AUROC for CURIAL-Rapide, assessed against PCR results. Further detail is provided in Appendix D.

## Results

### Patient Populations

Our updated training set (Supplementary Figure S2) comprised 114,957 patient presentations to OUH prior to the global COVID-19 outbreak (November 30, 2017 - December 01, 2019), considered as COVID-19-free presentations, and 701 patient presentations during the first wave of the UK COVID-19 epidemic (December 01, 2019 - June 30, 2021) who had a positive PCR test for COVID-19.

72,223 patients were included across four validation cohorts, of whom 4,600 had a positive confirmatory test for COVID-19 (Appendices A & B). In the external cohorts, patients admitted to PUH and BH trusts were similar ages (69 years (IQR 34) and 68 years (34); Kruskal-Walls p=0.9448), however patients admitted to UHB were significantly younger (63 years (37); p<0.0001 & <0.0001). A higher proportion of patients admitted to UHB were female (53.1%) than PUH and BH (45.0% and 46.7%; chi-square p<0.0001) and reported a South Asian ethnicity (13.2% versus 0.5% and 2.0%; chi-square p<0.0001).

Prevalence of COVID-19 was higher in the Bedfordshire cohort owing to the evaluation period matching the timeline of the second wave of the UK COVID-19 epidemic (11.1% versus 5.29% (PUH) and 4.27 (UHB); Fisher’s exact test p<0.0001 & <0.0001). Summaries of vital signs, index routine blood tests and blood gases are presented in Supplementary Tables S2-S4.

### Prospective evaluation of CURIAL-Lab & CURIAL-Rapide

Of 37,304 patients attending emergency departments and acute medical services across OUH during the second-wave of the COVID-19 pandemic (October 01, 2020 - March 06, 2021; Table 2 & Supplementary Figure S2), 14,447 were excluded as they did not receive confirmatory testing for COVID-19. We evaluated CURIAL-Lab and CURIAL-Rapide for all 22,857 patients receiving confirmatory testing (2056 testing positive), benchmarking performance against CURIAL-1.0 (Table 3). At the 80% sensitivity configuration, CURIAL-Lab performed similarly to CURIAL-1.0 (sensitives 72.9% & 73.6% respectively, and specificities 87.3% & 86.6%; McNemar chi-square test p=0.0823) but better than CURIAL-Rapide (sensitivity 74.7%, specificity 78.6%; p <0.0001) representing a trade-off between result-time and performance. CURIAL-Rapide and CURIAL-Lab achieved high NPVs (>98%) across both sensitivity configurations, achieving an AUROC of 0.843 and 0.878 respectively.

**Table 2:**
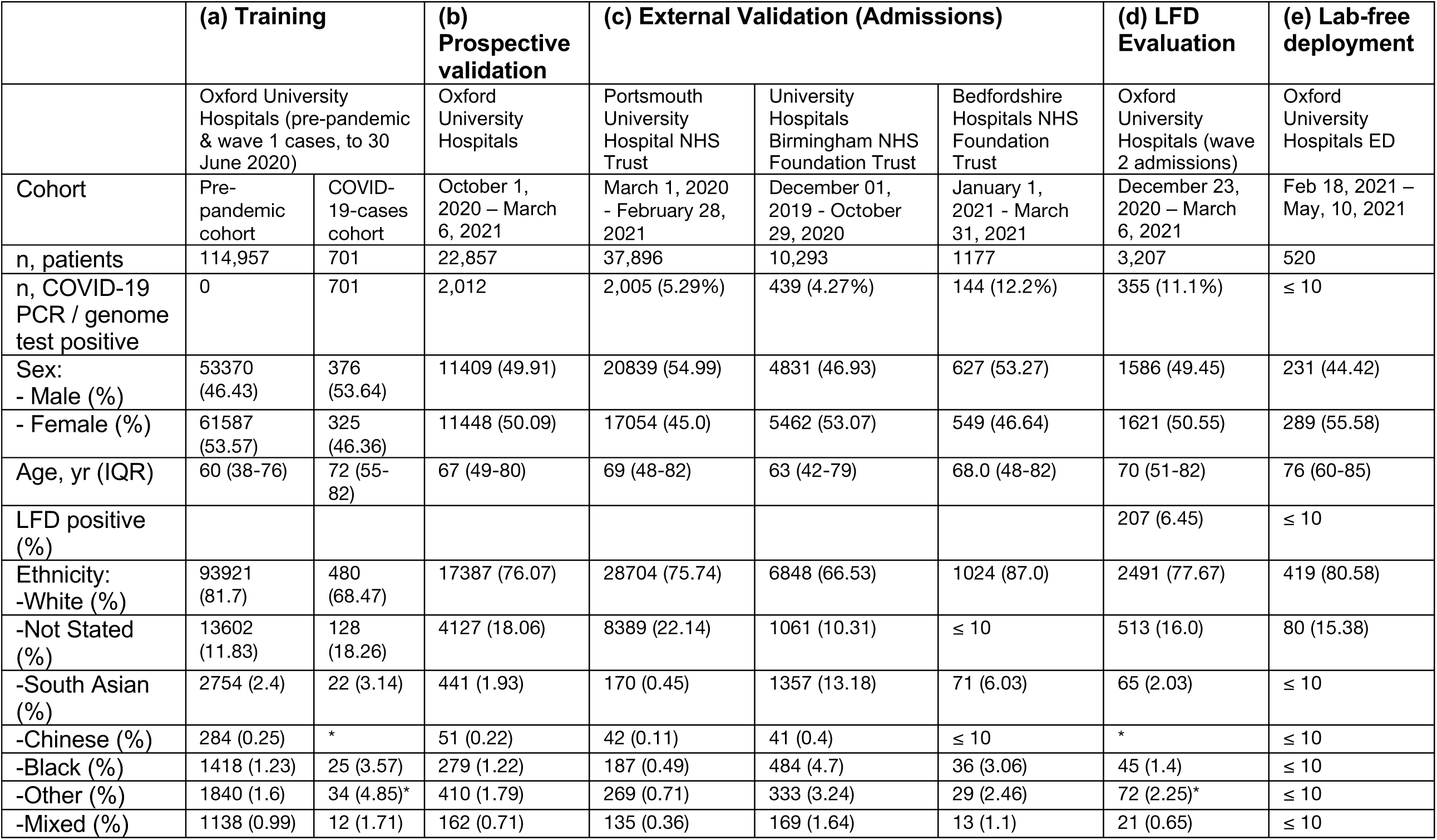
Summary population characteristics for (a) OUH pre-pandemic and COVID-19-cases training cohorts, (b) prospective validation cohort of patients attending OUH during the second wave of the UK COVID-19 epidemic, (c) independent validation cohorts of patients admitted to three independent NHS Trusts, (d) admissions to OUH during the second-wave receiving LFD testing, (e) patients enrolled to the CURIAL-Rapide lab-free service evaluation at OUH. The derivation of OUH cohorts is shown in Supplementary Figure S2. * indicates merging for statistical disclosure control.

|  | (a) Training |  | (b) Prospective validation | (c) External Validation (Admissions) |  |  | (d) LFD Evaluation | (e) Lab-free deployment |
| --- | --- | --- | --- | --- | --- | --- | --- | --- |
|  | Oxford University Hospitals (pre-pandemic & wave 1 cases, to 30 June 2020) |  | Oxford University Hospitals | Portsmouth University Hospital NHS Trust | University Hospitals Birmingham NHS Foundation Trust | Bedfordshire Hospitals NHS Foundation Trust | Oxford University Hospitals (wave 2 admissions) | Oxford University Hospitals ED |
| Cohort | Pre-pandemic cohort | COVID-19-cases cohort | October 1, 2020 – March 6, 2021 | March 1, 2020 – February 28, 2021 | December 01, 2019 – October 29, 2020 | January 1, 2021 – March 31, 2021 | December 23, 2020 – March 6, 2021 | Feb 18, 2021 – May, 10, 2021 |
| n, patients | 114,957 | 701 | 22,857 | 37,896 | 10,293 | 1177 | 3,207 | 520 |
| n, COVID-19 PCR / genome test positive | 0 | 701 | 2,012 | 2,005 (5.29%) | 439 (4.27%) | 144 (12.2%) | 355 (11.1%) | ≤ 10 |
| Sex: |  |  |  |  |  |  |  |  |
| - Male (%) | 53370 (46.43) | 376 (53.64) | 11409 (49.91) | 20839 (54.99) | 4831 (46.93) | 627 (53.27) | 1586 (49.45) | 231 (44.42) |
| - Female (%) | 61587 (53.57) | 325 (46.36) | 11448 (50.09) | 17054 (45.0) | 5462 (53.07) | 549 (46.64) | 1621 (50.55) | 289 (55.58) |
| Age, yr (IQR) | 60 (38-76) | 72 (55-82) | 67 (49-80) | 69 (48-82) | 63 (42-79) | 68.0 (48-82) | 70 (51-82) | 76 (60-85) |
| LFD positive (%) |  |  |  |  |  |  | 207 (6.45) | ≤ 10 |
| Ethnicity: |  |  |  |  |  |  |  |  |
| -White (%) | 93921 (81.7) | 480 (68.47) | 17387 (76.07) | 28704 (75.74) | 6848 (66.53) | 1024 (87.0) | 2491 (77.67) | 419 (80.58) |
| -Not Stated (%) | 13602 (11.83) | 128 (18.26) | 4127 (18.06) | 8389 (22.14) | 1061 (10.31) | ≤ 10 | 513 (16.0) | 80 (15.38) |
| -South Asian (%) | 2754 (2.4) | 22 (3.14) | 441 (1.93) | 170 (0.45) | 1357 (13.18) | 71 (6.03) | 65 (2.03) | ≤ 10 |
| -Chinese (%) | 284 (0.25) | * | 51 (0.22) | 42 (0.11) | 41 (0.4) | ≤ 10 | * | ≤ 10 |
| -Black (%) | 1418 (1.23) | 25 (3.57) | 279 (1.22) | 187 (0.49) | 484 (4.7) | 36 (3.06) | 45 (1.4) | ≤ 10 |
| -Other (%) | 1840 (1.6) | 34 (4.85)* | 410 (1.79) | 269 (0.71) | 333 (3.24) | 29 (2.46) | 72 (2.25)* | ≤ 10 |
| -Mixed (%) | 1138 (0.99) | 12 (1.71) | 162 (0.71) | 135 (0.36) | 169 (1.64) | 13 (1.1) | 21 (0.65) | ≤ 10 |

**Table 3:**
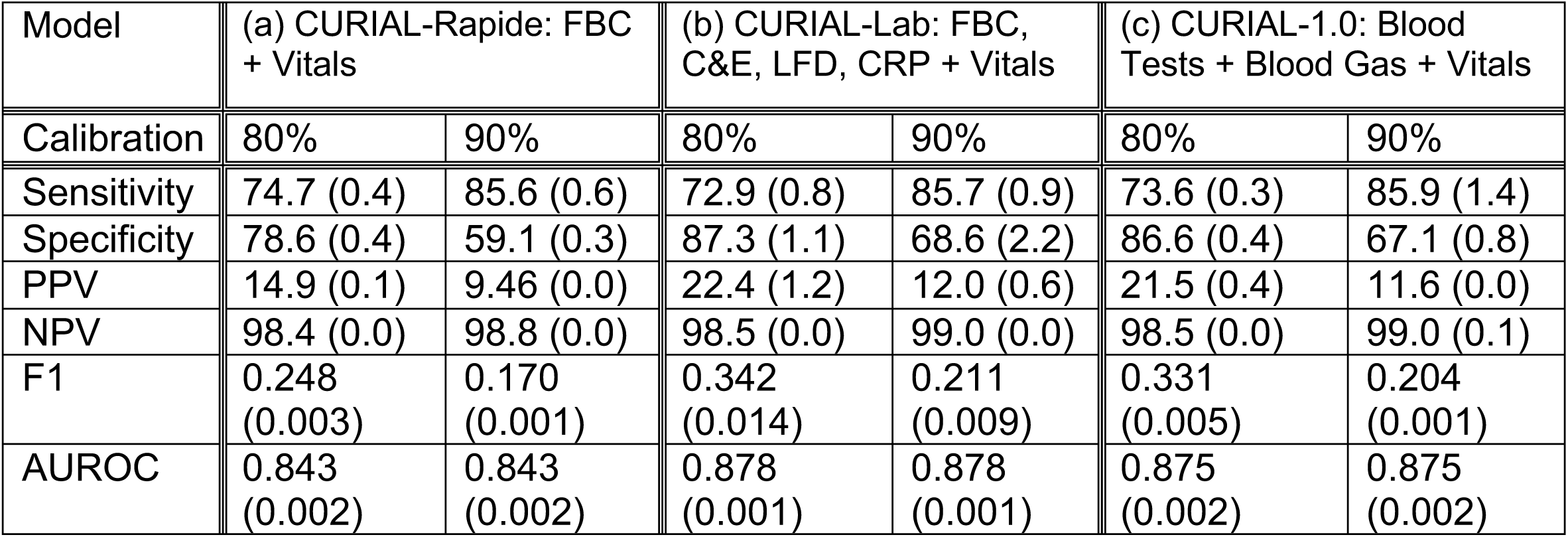
Evaluation of the performance of (a) CURIAL-Rapide and (b) CURIAL-Lab, calibrated during training to achieve sensitivities of 80% and 90%, on an independent prospective set of all admissions to OUH during the second-wave of COVID-19, between October 1, 2020 and March 6, 2021. (c) Benchmark performance of CURIAL-1.0 (Soltan et al. 2020) on the prospective set. Mean values are reported alongside SD across three imputation methods.

| Model | (a) CURIAL-Rapide: FBC + Vitals |  | (b) CURIAL-Lab: FBC, C&E, LFD, CRP + Vitals |  | (c) CURIAL-1.0: Blood Tests + Blood Gas + Vitals |  |
| --- | --- | --- | --- | --- | --- | --- |
| Calibration | 80% | 90% | 80% | 90% | 80% | 90% |
| Sensitivity | 74.7 (0.4) | 85.6 (0.6) | 72.9 (0.8) | 85.7 (0.9) | 73.6 (0.3) | 85.9 (1.4) |
| Specificity | 78.6 (0.4) | 59.1 (0.3) | 87.3 (1.1) | 68.6 (2.2) | 86.6 (0.4) | 67.1 (0.8) |
| PPV | 14.9 (0.1) | 9.46 (0.0) | 22.4 (1.2) | 12.0 (0.6) | 21.5 (0.4) | 11.6 (0.0) |
| NPV | 98.4 (0.0) | 98.8 (0.0) | 98.5 (0.0) | 99.0 (0.0) | 98.5 (0.0) | 99.0 (0.1) |
| F1 | 0.248 (0.003) | 0.170 (0.001) | 0.342 (0.014) | 0.211 (0.009) | 0.331 (0.005) | 0.204 (0.001) |
| AUROC | 0.843 (0.002) | 0.843 (0.002) | 0.878 (0.001) | 0.878 (0.001) | 0.875 (0.002) | 0.875 (0.002) |

A sensitivity analysis to assess susceptibility of our models to imputation strategy demonstrated consistent performance across multiple imputations (Table 3). We therefore performed subsequent evaluation using a model trained solely using a single imputation strategy (population median).

### External validation of CURIAL-Rapide, CURIAL-Lab and CURIAL-1.0

We externally validated CURIAL-Rapide & CURIAL-Lab at three independent hospital groups, across cohorts comprising 49,366 patient admissions (Figure 2, Supplementary Table S5). Performance was consistent across the trusts, with CURIAL-Lab achieving higher performance (AUROC range 0.858 - 0.881, 95% CIs range 0.838-0.912) than CURIAL-Rapide (AUROC range 0.836 - 0.854, 95% CIs range 0.814-0.889). Sensitivity of both models was higher when applied at Portsmouth University Hospitals (84.1%, 95% CI 82.5-85.7 & 83.5%, 81.8 - 85.1) compared to Bedfordshire Hospitals (74.3%, 66.6 - 80.7 & 74.3%, 66.6 - 80.7), at the expense of specificity (71.3%, 70.9 - 71.8 & 63.6%, 63.1 - 64.1 versus 84.8%, 82.5 - 86.9 and 81.8%, 79.3 - 84.0) possibly reflecting differences in confirmatory testing method at Bedfordshire Hospitals (SAMBA-II/Panther).

**Figure 2:**
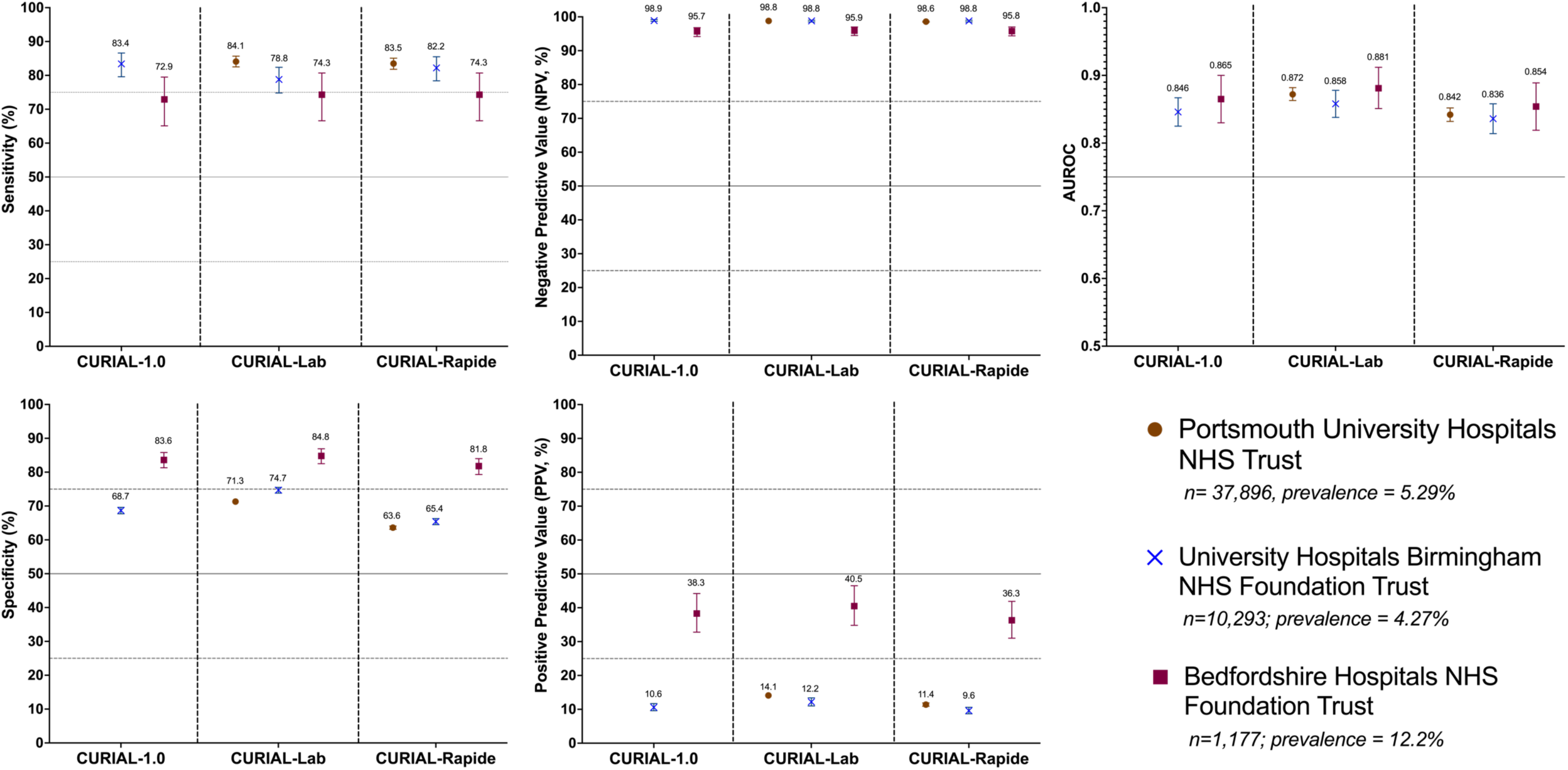
Performance of CURIAL-1.0, CURIAL-Lab & CURIAL-Rapide during external validation at three independent UK Hospitals trusts. All models were calibrated during training to achieve 90% sensitivity. Error bars show 95% confidence intervals. Numerical results are shown in Supplementary Table S5.

Both CURIAL-Rapide & CURIAL-Lab achieved high NPV across the three trusts in a prevalence-dependent fashion, with highest NPV at UHB where prevalence was lowest (4.27%, CURIAL-Rapide: 98.8% [98.5 - 99.0], CURIAL-Lab: 98.8% [98.5 - 99.0]). NPV was comparable at PUH where prevalence was similar (5.29%; CURIAL-Rapide: NPV 98.6% [98.4 - 98.7], CURIAL-Lab: 98.8% [98.6 - 98.9]). We additionally externally validated CURIAL-1.0 for the two trusts which electronically recorded blood-gas results, demonstrating comparable performance to CURIAL-Lab (Supplementary Table S5).

### Comparing CURIAL triage performance with LFDs & evaluating a combined CURIAL-LFD clinical pathway to enhance LFD sensitivity

We applied CURIAL-Rapide and CURIAL-Lab to the first-performed blood tests and vital signs for 3,207 patients admitted to OUH, receiving LFD testing from introduction on December 23, 2020 to March 6, 2021 (Figure 3, Supplementary Table S6). Patient eligibility is demonstrated in Supplementary Figure S2. One patient with an invalid LFD result was excluded from analysis. LFDs achieved a high specificity of 99.8% (99.6 - 99.9) during triage, but poor sensitivity of 56.9% (51.7 - 62.0).

**Figure 3:**
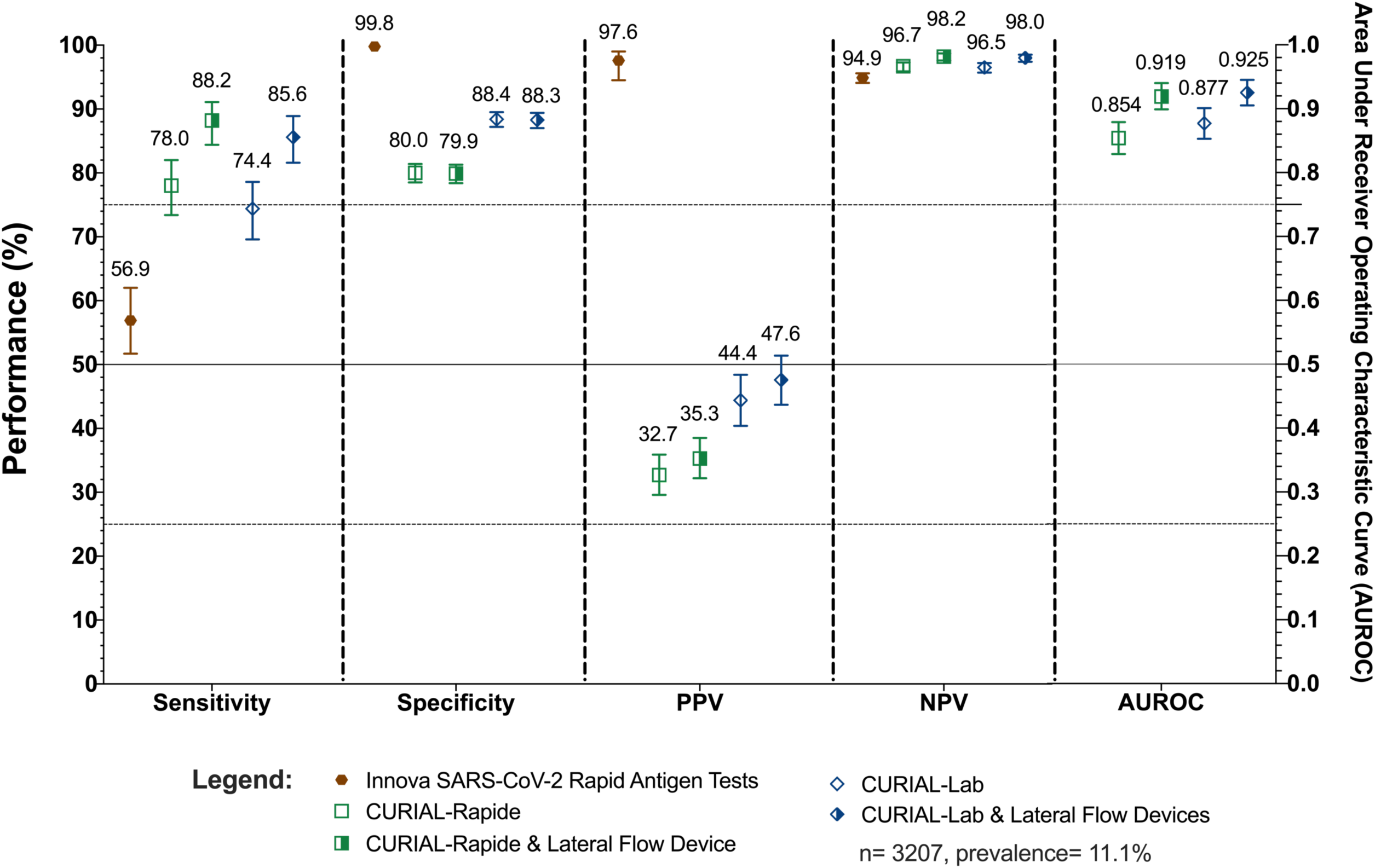
Performance characteristics of (a) INNOVA SARS-CoV2 Rapid Antigen Tests, (b) CURIAL-Rapide & CURIAL-Lab, calibrated during training to a sensitivity of 80%, and (c) combined clinical pathways considering either a positive CURIAL-Rapide/CURIAL-Lab result or a positive LFD test as a COVID-19 suspected case, at Oxford University Hospitals NHS Foundation Trust between December 23, 2020 & March 6, 2021. Error bars show 95% confidence intervals. Numerical results are shown in Supplementary Table S6.

CURIAL-Rapide & CURIAL-Lab were significantly more sensitive (78.0% [73.4 - 82.0] & 74.4% [69.9 - 78.6]) than LFDs, therefore achieving higher negative predictive values (CURIAL-Rapide: 96.7% [95.9-97.3], CURIAL-Lab: 96.5% [95.7 - 97.2], versus LFDs: 94.9% [94.1 - 95.6]). By contrast, the models were less specific (80.0% [78.5 - 81.4] & 88.4% [87.2 - 89.5]), thereby favouring higher reliability for safely excluding COVID-19 at the expense of the false positive rate.

Integrating positive LFD results with CURIAL-Rapide/CURIAL-Lab, as part of a combined clinical pathway (Figure 1), significantly improved overall sensitivity (to 88.2% [84.4 - 91.1] & 85.6% [81.6 - 88.9]) and NPV (to 98.2% [97.6 - 98.7] & 98.0% [97.4 - 98.5]), reducing COVID-19 status misclassification (McNemar’s chi-square test, p=0.0003 & p=0.0004). AUROC was significantly improved to 0.919 (0.899 - 0.940) for a CURIAL-Rapide/LFD pathway, and 0.925 (0.905 – 0.945) for CURIAL-Lab/LFD pathway. Performance of CURIAL-Lab was similar to CURIAL-1.0 (p=0.860; Supplementary table S6).

### Deployment & operational evaluation of CURIAL-Rapide at Oxford University Hospitals NHS Foundation Trust

520 patients were enrolled to the OLO/CURIAL-Rapide service evaluation between February 18, 2021 and May 10, 2021 (baseline characteristics, Table 2). 436 patients received confirmatory PCR testing within routine care, of whom 10 returned positive results (prevalence 2.3%). This reflected the falling prevalence of COVID-19 in the UK, coinciding with governmental restrictions and the national vaccination programme^32^. 348 patients received LFDs within routine care, with 4 positive results. Two patients with indeterminate PCR results were excluded from analysis, although both had a negative LFD result and were triaged by the assessing clinician to a COVID-19-free clinical pathway. A summary of OLO results and vital signs are shown in Supplementary Tables S2-S3.

Median time from registration in the ED to availability of a CURIAL-Rapide result was 45:00 minutes (32:00-64:00), 16 minutes (26.3%) sooner than for LFDs (61:00 min, IQR 36:45-99:00; Wilcoxon Signed Rank test p<0.0001), and 6h 52 minutes (90.2%) sooner than RT-PCR results (07:37 h, IQR 06:05-15:39; p<0.0001). Kaplan Meier survival analyses (Figure 4a) showed CURIAL-Rapide results were available sooner than LFDs (log rank test, p<0.0001) and PCR results (p<0.0001). The median time-to-result for full blood count analysis was shorter with near-patient OLO analysis (44:00 min, 31:00-63:00 min) than laboratory analysis (76:00 min, 58:00-100:00 mins; p <0.0001), confirming an improvement to routine care.

**Figure 4:**
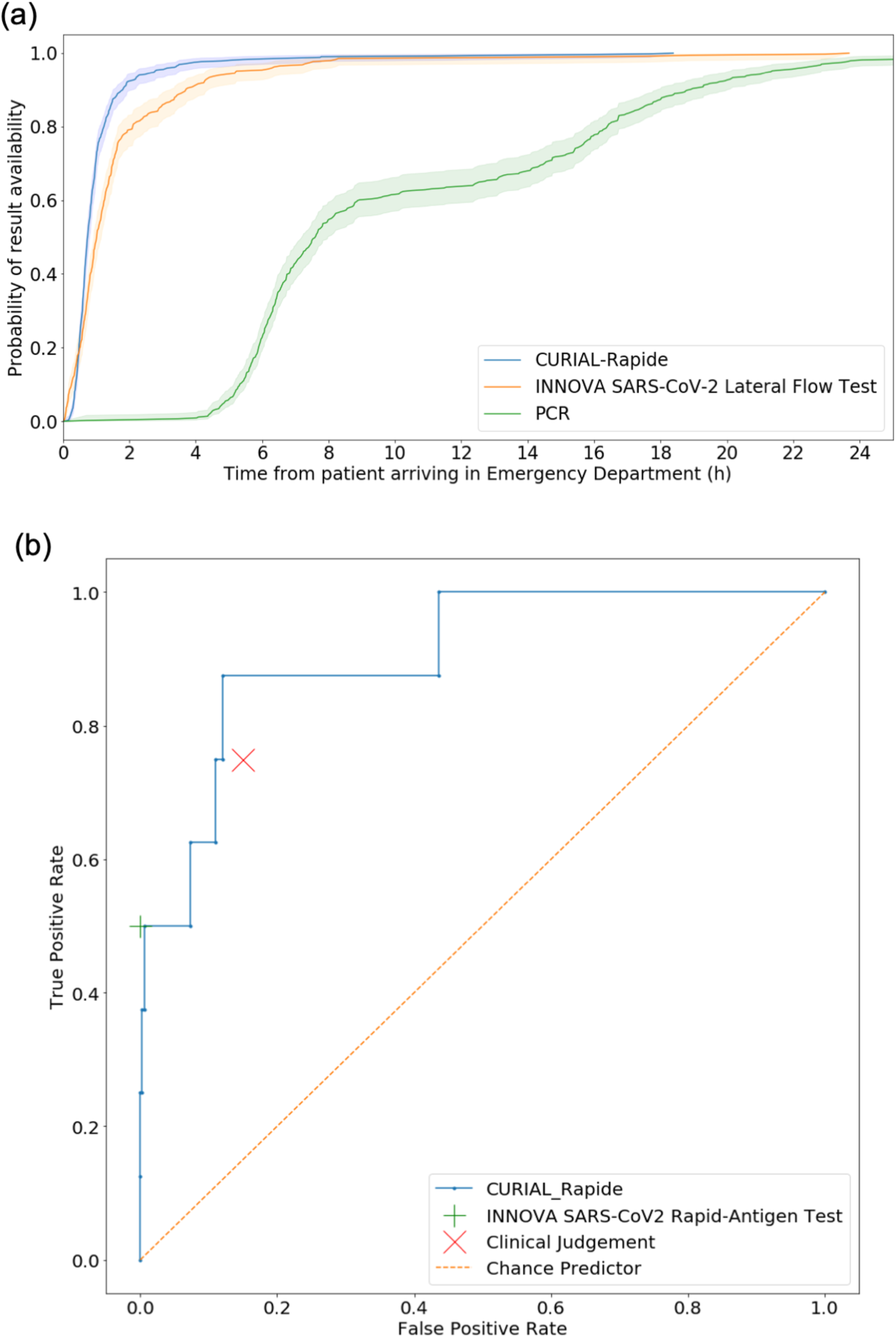
(a) Kaplan-Meier plots of time-to-result in hours from patient arrival in the Emergency Department for (i) CURIAL-Rapide, (ii) INNOVA SARS-CoV-2 Rapid-Antigen Tests and (iii) PCR swabs tests (Numbers at risk: 520, 348 and 436 respectively). CURIAL-Rapide results were available sooner than LFD testing (log rank test, p<0.0001) and PCR test results (p<0.001). (b) Receiver operating characteristic curve showing performance of (i) CURIAL-Rapide (ii) clinical judgement of the first-attending clinician, and (iii) INNOVA SARS-CoV-2 rapid antigen testing, against a PCR reference standard.

CURIAL-Rapide results had a negative predictive value of 99.7% (98.2-99.9), specificity of 85.4% (81.3-88.7), and AUROC of 0.907 (0.803-1.00). The point estimate of CURIAL-Rapide’s sensitivity was 87.5%, however 95% CIs were wide owing to the lower-than-expected prevalence of COVID-19 (52.9-97.8)).

In one presentation, a patient given a ‘negative’ CURIAL-Rapide prediction went on to have a positive SARS-CoV-2 PCR test, although they had a negative LFD result and were triaged to a COVID-19-free (‘green’) clinical area. The patient did not have COVID-19 symptoms on this presentation. We noted that the patient had also been enrolled to the service evaluation 10 days prior; on that occasion having a positive CURIAL-Rapide prediction, positive Lateral Flow Test and a positive PCR test. This raises the possibility of a latent positive PCR result, detecting non-infectious residual viral fragments, on the date of the second presentation^39,40^.

Rates of COVID-19 status misclassification were comparable between CURIAL-Rapide and clinician judgement (McNemar’s Exact test; p=0.91). Moreover, of the 53 patients who were triaged to a ‘COVID-19-suspected’ (amber) pathway by the attending clinician but went on to test negative by PCR, 31 patients (58.5%) had a negative CURIAL-Rapide prediction demonstrating that the AI system could reduce operational strain by expediting clinical exclusion of infection. As all patients with positive LFD results also had positive CURIAL-Rapide predictions, a combined CURIAL-Rapide/LFD pathway did not impact classifier performance in this evaluation.

## Discussion

National health policy recognises that effective in-hospital triage is necessary to safeguard patient and staff safety, mandating COVID-19 testing for all admissions^4^. However, despite significant innovation leading to new near-patient testing options, alongside reduced PCR result-times to typically within 12-24 h, there remain significant performance and logistical limitations that contribute to nosocomial transmission and operational strain. While many hospitals have adopted LFDs within acute admissions pathways^10^, our study confirms a limited sensitivity (56.9%) indicating a clinically-meaningful false negative rate (Figure 3, Supplementary Table S6)^14–16,41^.

In this study we demonstrate generalisability, efficacy, and real-world operational benefits of AI-driven COVID-19 screening in the acute care setting. Whereas rapid molecular testing options are frequently rationed^11,12^, we show that a high-throughput AI-solution, CURIAL-Lab, rapidly excludes COVID-19 using routine data and generalises across three independent hospital groups (Figure 2). Moreover, we improve upon the speed of existing rapid testing solutions, demonstrating a median result-time of 45 minutes (32-64 min; CURIAL-Rapide) from patients’ first arrival in an emergency department using near-patient haematology analysis. This decentralised approach may support time-critical decision making and assist triage in remote and primary care settings where laboratory facilities are less readily available.

In our external and prospective validation of CURIAL-Lab & CURIAL-Rapide, model performance was consistently high across four UK hospital groups (CURIAL-Lab: AUROC range 0.858-0.881; CURIAL-Rapide 0.836-0.854), with high negative predictive values confirming suitability as tests-of-exclusion for COVID-19. CURIAL-Lab expectedly achieved marginally superior performance to CURIAL-Rapide, representing a trade-off between result-time and specificity which would favour different clinical use-cases. Strengths of the validation include geographic breadth, including over 72,000 patients across three regions of the UK (Midlands, South East, and East of England), and temporal breadth across both waves of the UK COVID-19 epidemic therefore including vaccinated patients and patients with Coronavirus variants, across a range of prevalences (4.27%-12.2%). Moreover, the validation study considers the broad range of confirmatory COVID-19 tests (PCR, SAMBA-II & Panther) utilised across different centres, and address sector-wide concerns highlighted in key reviews of COVID-19 diagnostic and prognostic models by using external, representative cohorts of all unscheduled adult admissions^27,28^. Notable limitations include that the external validation is solely UK-based and the limitations of the confirmatory testing methods, with PCR testing having been shown to be imperfectly sensitive^6,7^. We were unable to quantify the number of vaccinated patients as we could not link our de-identified hospital datasets with vaccination records.

Our study finds that CURIAL-Rapide & CURIAL-Lab achieve significantly higher sensitivity and NPV than LFDs, improving upon standard-care by reducing risk of a COVID-19-positive patient being streamed to a COVID-19-free clinical area. Moreover, our study is the first to validate an application of an AI test to enhance sensitivity of LFDs in a real-world clinical setting, with our combined clinical pathway (Figure 1; CURIAL-Lab/LFD) achieving both high sensitivity of 85.6% (81.6 - 88.9) and overall classification performance with AUROC of 0.925 (0.905 - 0.945). A significant beneficiary population includes patients streamed to COVID-19-free (‘green’) clinical areas on receiving negative LFD and CURIAL results, which are performed in parallel and available much sooner than PCR results. Moreover, our pathway identifies an enriched subpopulation at greater risk of testing positive for COVID-19 (negative LFD & positive CURIAL-Lab/Rapide compared to negative LFD alone), therefore enabling prioritisation for rapid confirmatory testing where availability is limited. Limitations of this analysis include that patients may have been inadvertently excluded if LFDs were recorded incorrectly on the EHR, and the analysis was performed only for a single trust as other participating sites did not electronically record LFD results.

We report the fastest result-time to date for AI-driven COVID-19 screening in a hospital emergency department, using lab-free haematology analysis to achieve a median reduction of 16 minutes (26.3%) over LFDs. Significantly, by demonstrating that CURIAL-Rapide correctly excluded COVID-19 for 58.5% of negative patients who were triaged by a clinician to a ‘COVID-19-suspected’ (amber) clinical area, we show a role for AI screening in reducing delays in transfers to wards. A strength of our service evaluation is its real-world context and operational focus, assessing time from first-arrival to result availability, and demonstrating added clinical value by comparison to LFD results and clinician impressions. In this study, we address the need for evidence of clinical utility that AI tools such as CURIAL offer to the pandemic response^30^; demonstrating both operational and safety improvements to standard-care.

A significant limitation of our service evaluation is that, although the a priori target sample size was achieved, the desired precision and power levels were not achieved for the metric of sensitivity due to sharply falling prevalence of COVID-19 in the UK, associated with the national vaccination programme and public health measures^32,42^. The evaluation was, however, adequate to determine specificity. As a service evaluation, we used a convenience series, with OLO operation limited to daytime and evening hours (8am-8pm) for logistical reasons. Moreover, although routine LFD testing was hospital policy, 33% of enrolled patients did not have a coded result in their EHR using the pre-specified form raising the possibility that these may have been recorded elsewhere or communicated verbally. As all patients who were LFD positive also had a positive CURIAL-Rapide result in our study, a larger evaluation is needed to assess whether integrating LFD results could further improve performance of CURIAL-Rapide in this context. Further evaluation would assess performance as a clinical decision support aid and for sensitivity across coronavirus variants.

A major strength of the CURIAL-Rapide and CURIAL-Lab solutions is the use of clinical data that is readily available and routinely collected for all patients admitted to hospital. Our approach optimises generalisability with CURIAL-Lab, applicable to virtually all unscheduled patient admissions to hospital, thereby facilitating COVID-19-screening without significant additional cost. Where faster exclusion of COVID-19 is helpful for operational or treatment reasons, CURIAL-Rapide can provide faster results by eliminating the need for blood sample transportation and laboratory processing, at an approximate cost of around ∼£9 (∼$12.50; inclusive of device rental and consumables). By contrast, alternative strategies for AI-assisted COVID-19 diagnosis largely focus on chest imaging^23,28,43^, which involve patient exposure to ionising radiation and have higher costs. Following successful COVID-19 vaccination programmes, falling community prevalence may reduce cost-effectiveness of universal PCR-testing for unscheduled admissions. Our results suggest that CURIAL-Lab could deliver significant cost savings by reducing the number of routine PCR tests by >85% (where prevalence <2%) while achieving high NPV, utilising data that would be collected within the routine course of a patients’ care.

Our work demonstrates generalisability, efficacy, and real-world operational benefits of AI-driven COVID-19 screening for patients attending hospital. Future work would assess international generalisability, evaluate clinician-model interactions, and assess sensitivity of model performance across vaccination types and infection with variants of concern.

## Supporting information

STARD Checklist

## Abbreviations

AI: Artificial Intelligence
AUROC: Area under receiver operating characteristic curve
BH: Bedfordshire Hospitals NHS Foundation Trust
COVID-19: Coronavirus Disease 2019
CRP: C-Reactive Protein
ED: Emergency Department
EHR: Electronic Health Records
FBC: Full Blood Count
LFT: Liver Function Test
LFD: Rapid antigen lateral flow device
NHS: National Health Service
NPV: Negative Predictive Value
OUH: Oxford University Hospitals NHS Foundation Trust
PCR: Real Time Polymerase Chain Reaction
POCT: Point of Care Test
PPV: Positive Predictive Value
PUH: Portsmouth University Hospitals NHS Trust
SARS-CoV-2: Severe Acute Respiratory Syndrome Coronavirus 2
SD: Standard Deviation
U&Es: Urea, Creatinine and Electrolytes
UHB: University Hospitals Birmingham NHS Foundation Trust

## Acknowledgments

We express our sincere thanks to all patients and staff across the four participating NHS trusts; Oxford University Hospitals, University Hospitals Birmingham, Bedfordshire Hospitals, and Portsmouth University Hospitals NHS trusts. We particularly wish to thank all nursing, medical and allied healthcare professional colleagues in the John Radcliffe Hospital’s Emergency Department for supporting evaluation of the OLO analyser and CURIAL-Rapide. We thank Emergency Medicine Research in OXford (EMROX), Dr Natasha Dole, Dr Jaspreet Bahra and Dr Ayisha Shaikh (Emergency Department Registrars), Ian Smith, Vikram Lyall and the OUH Point-of-care testing office. We additionally express our gratitude to Jingye Wang & Jolene Atia at University Hospitals Birmingham NHS Foundation trust, Phillip Dickson at Bedfordshire University Hospitals, and Paul Meredith at Portsmouth University Hospitals for assistance with data extraction.

## Declarations

DWE reports personal fees from Gilead, outside the submitted work; DAC reports personal fees from Oxford University Innovation, personal fees from BioBeats, personal fees from Sensyne Health, outside the submitted work. No other authors report any conflicts of interest.

## Contributions

AS, AN, RP, MS, DRT, TZ, DWE & DAC conceived of and designed the study. DWE, RF, AN, RP, MS, DRT, SB & CURIAL Translational Collaborative assisted with data collection/extraction and study operation. AS, JY, YY, OR pre-processed and verified the data, and performed the analyses. AS and DWE wrote the manuscript. All authors revised the manuscript.

## Ethics

NHS Health Research Authority (HRA) approval was granted for the use of routine clinical and microbiology data from Electronic Health Records for development and validation of artificial intelligence models to detect Covid-19 (CURIAL; NHS HRA IRAS ID 281832).

## Data & Code Availability

Data from OUH studied here are available from the Infections in Oxfordshire Research Database (https://oxfordbrc.nihr.ac.uk/research-themes-overview/antimicrobial-resistance-and-modernising-microbiology/infections-in-oxfordshire-research-database-iord/), subject to an application meeting the ethical and governance requirements of the Database. Data from UHB, PUH and BH are available on reasonable request to the respective trusts, subject to HRA requirements. Code and supplementary information for this paper are available online alongside publication.

## Supplementary Material

## Appendix A CURIAL-Rapide Translational Collaborative

We thank all healthcare professionals and students who have supported the CURIAL-Rapide Service evaluation. In particular, we wish to acknowledge:

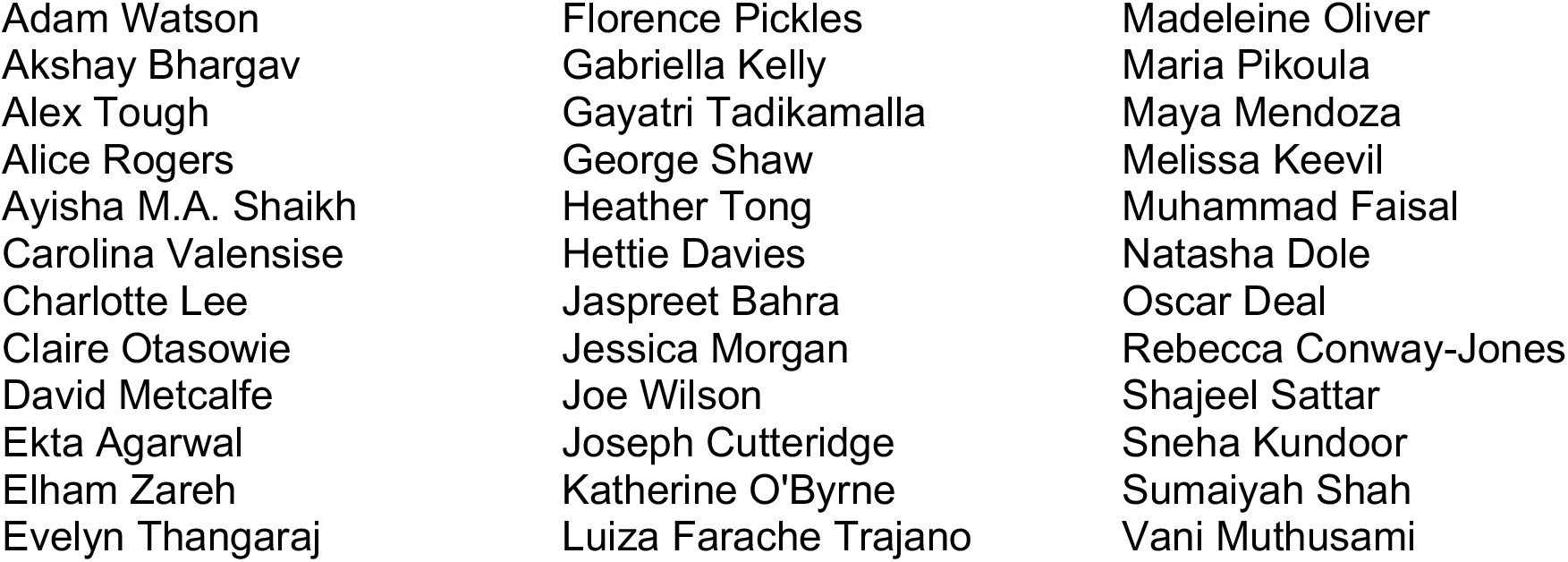

## Appendix B

**Supplementary Figure S1:**
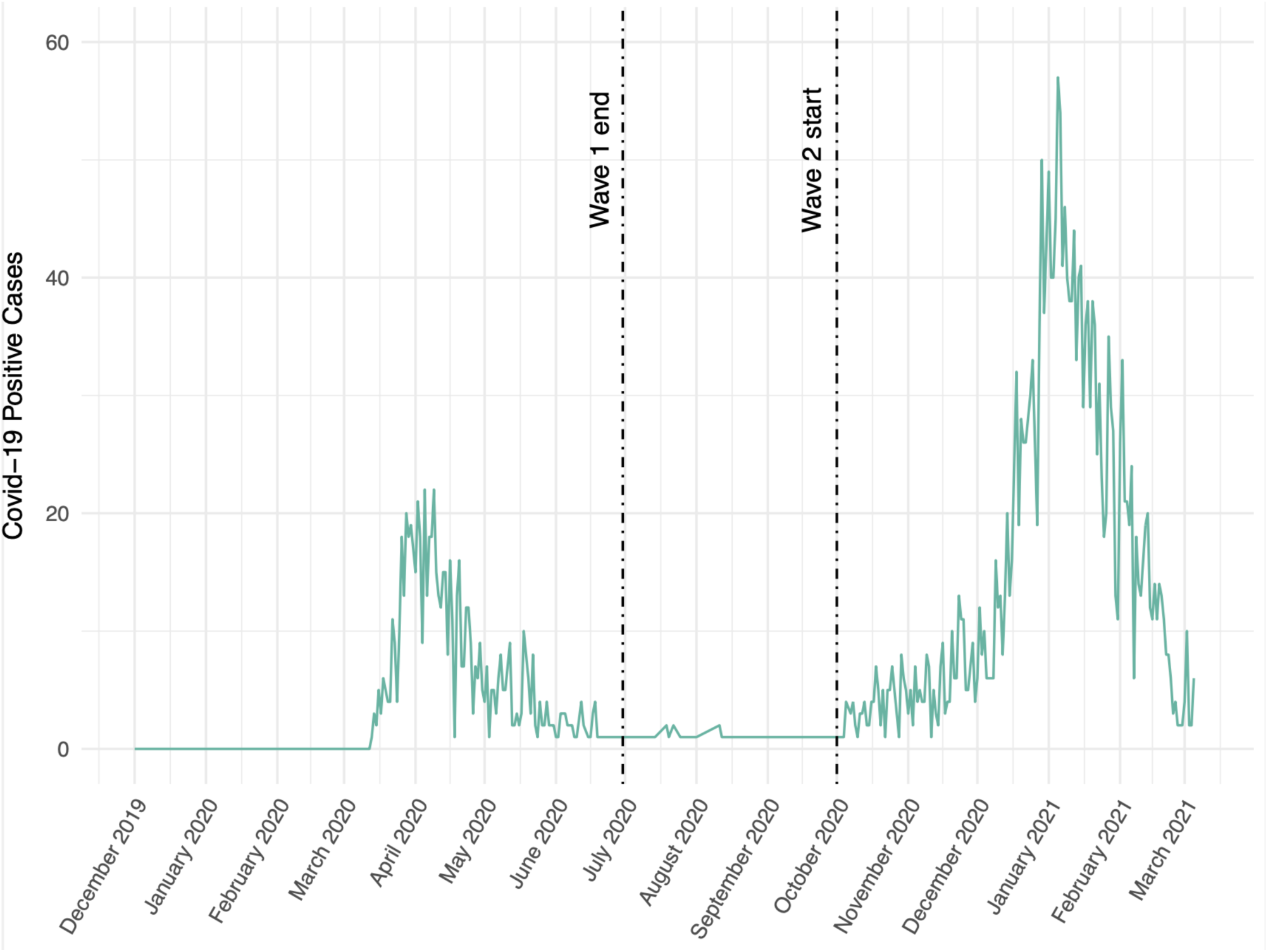
Daily number of patients presenting to Oxford University Hospitals NHS Foundation Trust testing positive for COVID-19, between 1^st^ December 2019 and 8^th^ March 2021.

### Model Development: Inclusion & Exclusion Criteria

As previously, we included all patients attending acute and emergency care settings at Oxford University Hospitals NHS foundation trust who received routine blood tests on arrival, considering presentations before December 1, 2019, and thus before the pandemic, as the COVID-19-negative (control) cohort. We considered presentations during the ‘first wave’ of the UK COVID-19 pandemic (December 1, 2019 to June 30, 2020) with PCR confirmed SARS-CoV-2 infection as the COVID-19-positive (cases) cohort. We excluded patients who opted out of electronic health record (EHR) research and those who did not receive laboratory blood tests or were younger than 18 years of age. Due to incomplete penetrance of testing during the first wave of the pandemic, and imperfect sensitivity of the PCR test, there is uncertainty in the viral status of patients presenting during the pandemic who were untested or tested negative. We therefore selected a pre-pandemic control cohort during training to ensure absence of disease in patients labelled as COVID-19-negative.

Clinical features extracted for each presentation included first-performed blood tests, blood gases, vital signs measurements and PCR testing for SARS-CoV-2 (Abbott Architect [Abbott, Maidenhead, UK], TaqPath [Thermo Fisher Scientific, Massachusetts, USA] and Public Health England-designed RNA-dependent RNA polymerase assays). A list of extracted clinical features is shown in Supplementary Table S1.

#### Normalisation

Data normalisation was implemented to mitigate overfitting and to avoid the reliance of the model on measurement units. Categorical data were handled by encoding as “1-hot” variables. Where a lab value was reported as being below the threshold of detection of the laboratory assay, the value was replaced with a numerical zero value. Where values were reported as being above the threshold of detection, clinically appropriate values were selected to maintain the significance of the high result. A summary of first-performed blood tests, vital signs, and blood gasses on arrival to hospital are shown Supplementary Tables S2-S4.

#### Missing Data

Multiple imputation strategies, population median, population mean, and age-based imputation, were separately used to impute missing data initially. As a sensitivity analysis to assess for effects of imputation strategy on model performance, we assessed performance of models trained using each imputation method prospectively for all patients attending emergency departments and acute medical services across OUH during the second-wave of the COVID-19 pandemic (October 01, 2020 and March 06, 2021; Table 2). Mean performance was reported alongside SD in Table 2, with narrow standard deviations in all performance metrics demonstrating resilience to imputation method. We therefore subsequently only used models trained with missing data imputed using population median, reporting results alongside 95% confidence intervals (CIs).

### Model Training & Prospective Evaluation

We repeated training and optimisation of our eXtreme Gradient BOOSTed tree model (XGBoost) to discriminate COVID-19-positive cases from pre-pandemic COVID-19-negative controls, for each of the three feature-sets (Table 1) ^44^. During training using ‘first wave’ case, controls were matched for age, gender, and ethnicity at a ratio of 1:20. For missing data, we initially used three independent imputation methods during training – median, mean and age-based mean – and assessed sensitivity of model performance to imputation strategy during testing. Thresholds were calibrated to achieve sensitivities of 80% and 90% during training, using stratified 10-fold cross validation.

XGBoost is a generalisation of boosting to an arbitrary differentiable loss function. XGBoost is more robust to outliers and has high predictive power. The scikit-learn (v0.23.2), LIBLINEAR (v2.41) and XGBoost (v1.2.0) modules for Python were used during model development and classifier evaluation.

**Supplementary Table S1:**
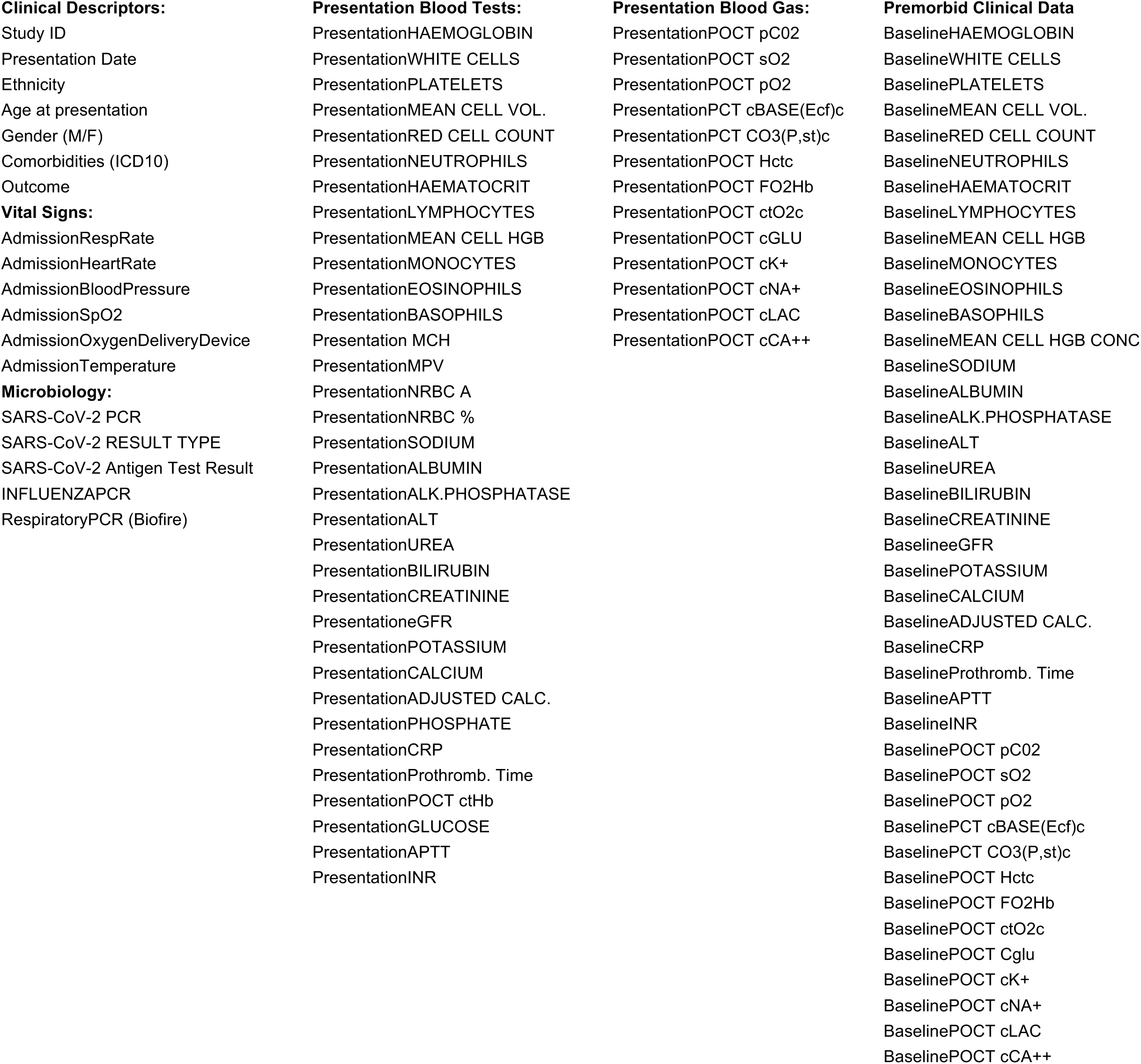
Clinical data fields extracted from training and externally validating NHS sites, for all patients admitted to the trusts from 1^st^ December 2019 onwards, and for up to 200,000 pre-pandemic admissions.

**Supplementary Figure S2:**
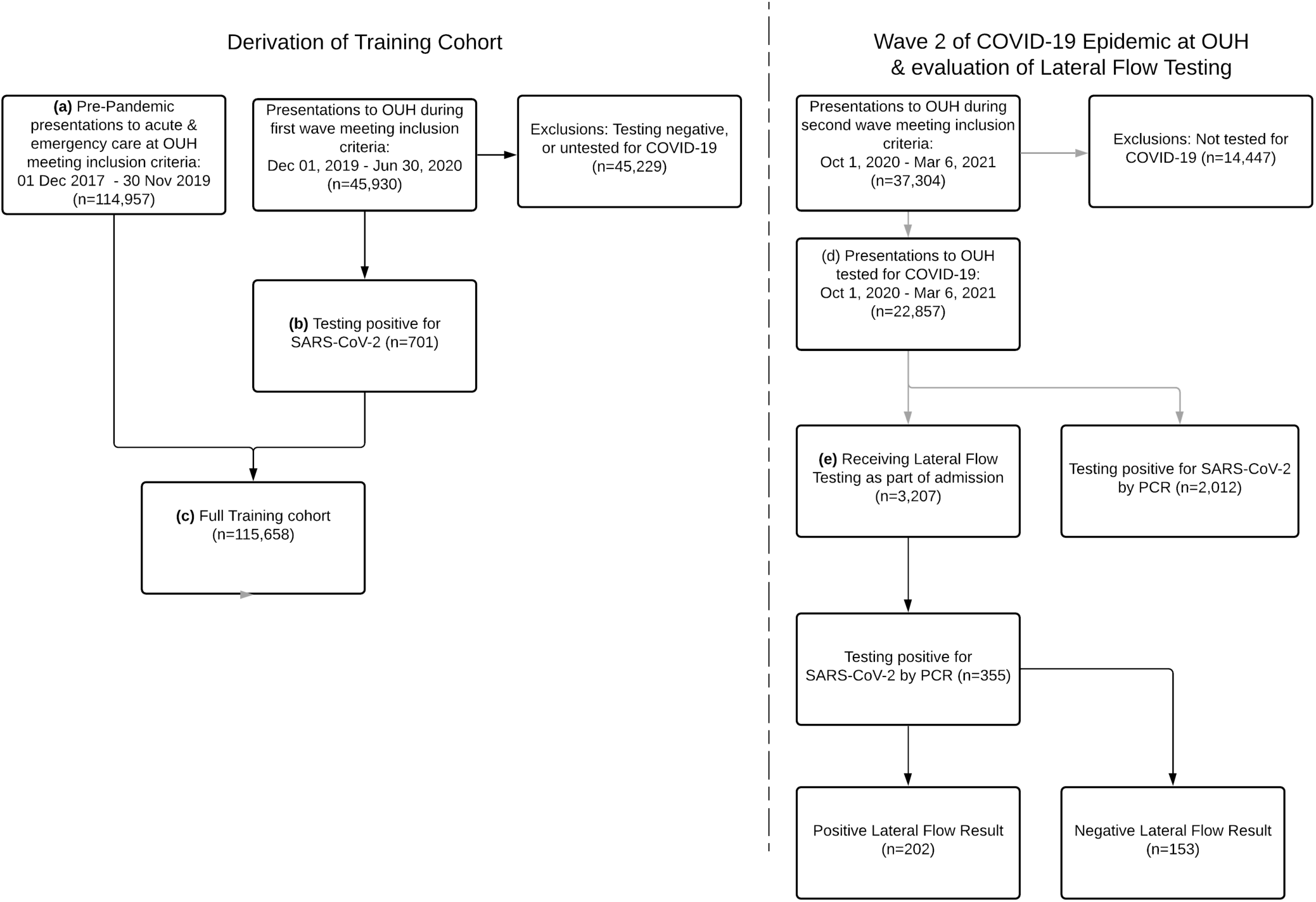
Participant flow diagram showing patients attending OUH, who met inclusion and exclusion criteria, for (a) the pre-pandemic training cohort and (b) COVID-19-cases cohort, combining to form (c) a full training cohort for model development. Patients attending OUH during the second wave of the UK COVID-19 epidemic, between Oct 1, 2020 and Mar 6, 2020, meeting inclusion and exclusion criteria, formed (d) the second wave analysis cohort, of which a subset (e) received Lateral Flow Testing within routine care, as part of an admission.

**Supplementary Table S2:**
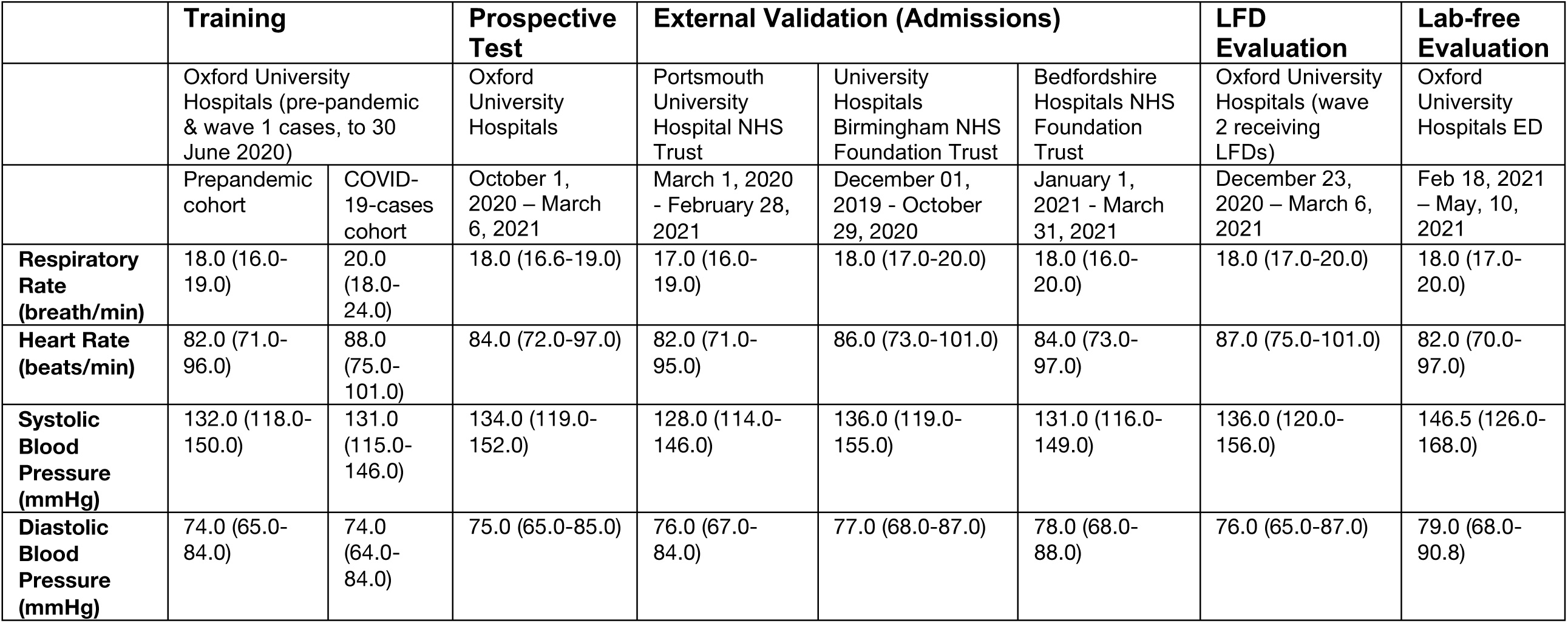
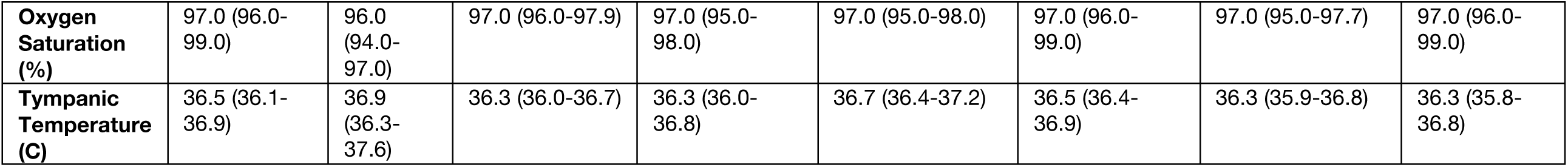
Distribution of vital signs, reported as median and interquartile ranges, for each patient cohort.

**Supplementary Table S3:**
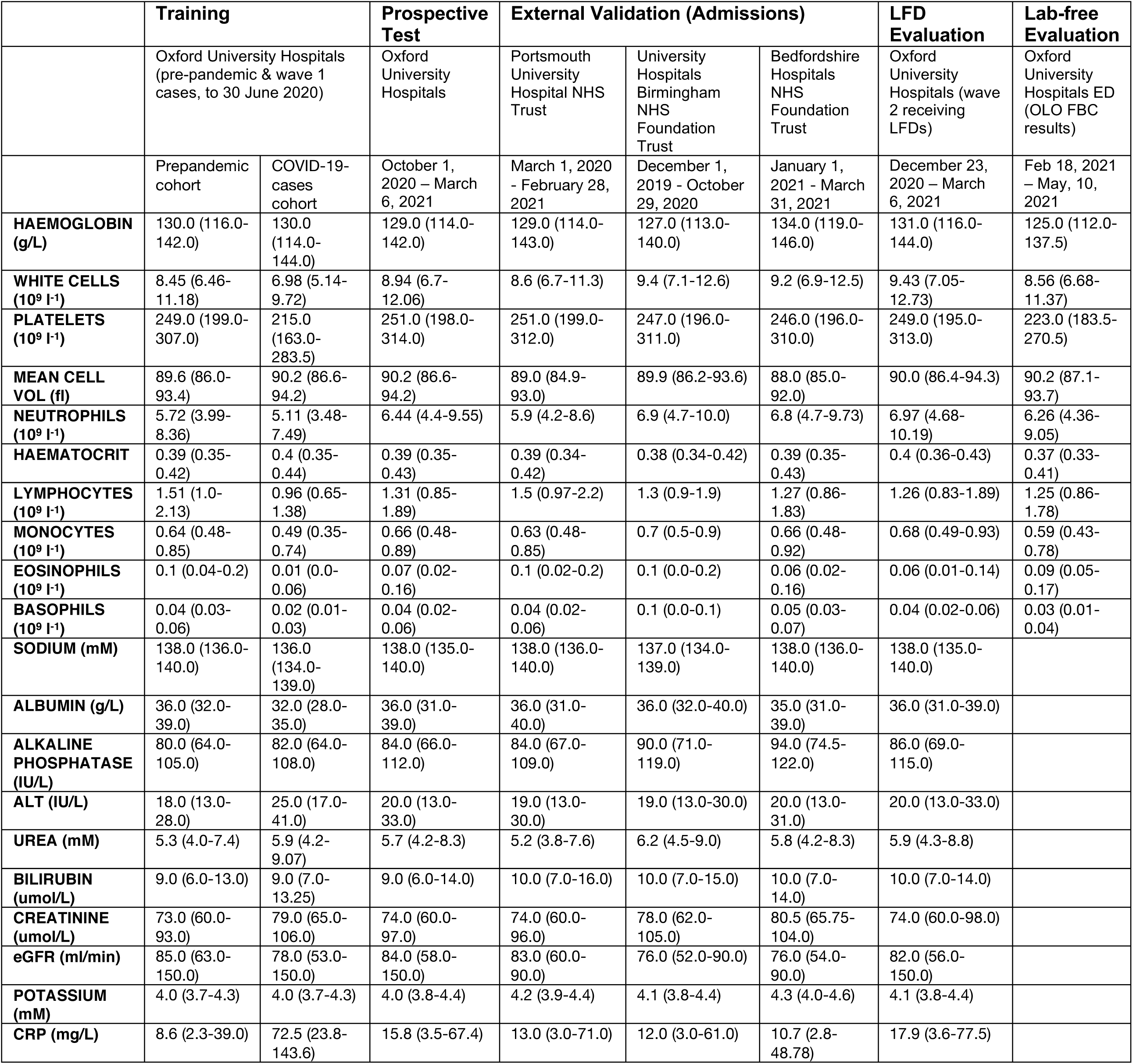
Distribution of blood test features, reported as median and interquartile ranges, for each patient cohort.

**Supplementary Table S4:**
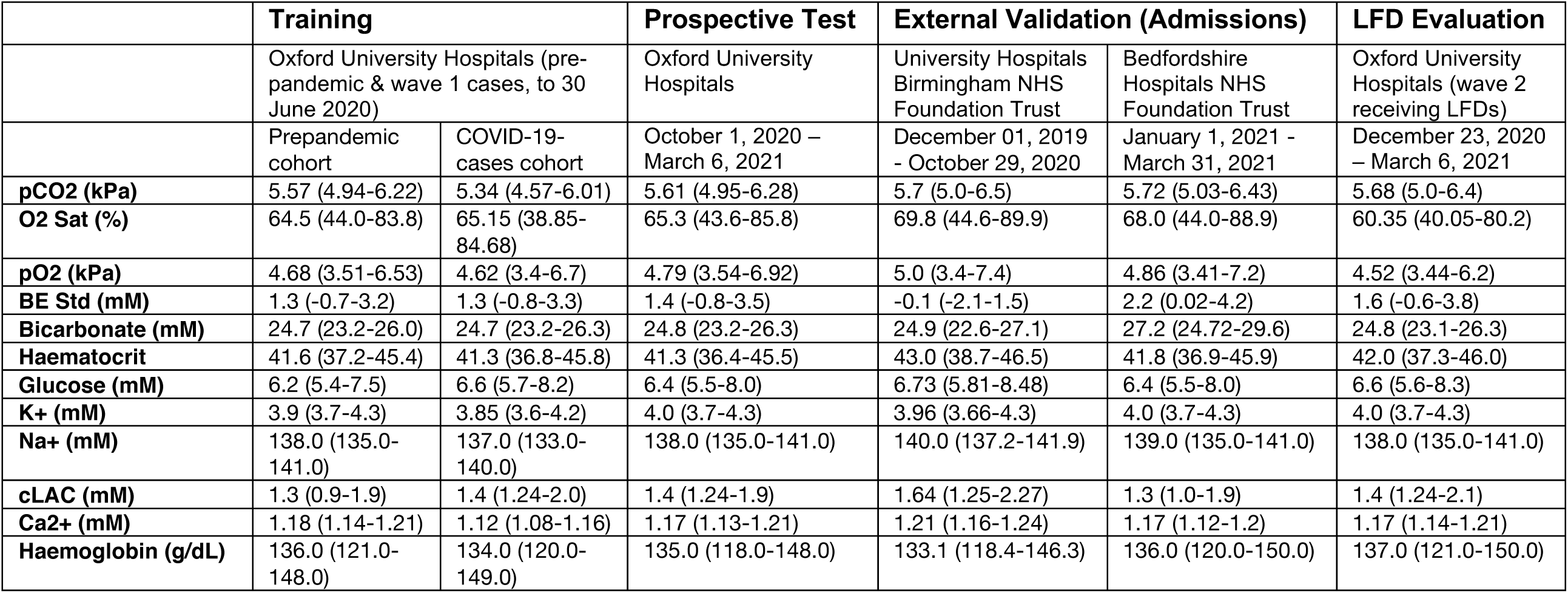
Distribution of blood gas features, reported as median and interquartile ranges for each patient cohort.

## Appendix C: External validation at independent NHS Trusts

We externally validated CURIAL-Rapide and CURIAL-Lab by applying the respective models to results of first-available blood test results and vital signs (Table 1), comparing model predictions to confirmatory SARS-CoV-2 viral genome test results. For trusts where blood-gas results were available for electronic extraction, we also evaluated CURIAL-1.0. Patients meeting inclusion criteria had an unscheduled acute or emergency care admission, during the specified periods, received a blood draw on arrival and were aged over 18. We excluded patients who did not have a valid confirmatory test result within a prespecified period, or who had opted out of EHR research. Screening against eligibility criteria, followed by anonymisation, was performed by the respective NHS Trusts.

Evaluation at Portsmouth Hospitals NHS Foundation Trust (PUH) considered all patients admitted to the Queen Alexandria Hospital, serving a population of 675,000 and offering tertiary referral services to the surrounding region, between March 1, 2020 and February 28, 2021. Confirmatory COVID-19 testing was by laboratory SARS-CoV-2 RT-PCR assay, considering any positive PCR result within 48hrs of admission as a true positive. As blood gas results were not available for electronic extraction, we evaluated only CURIAL-Rapide and CURIAL-Lab at Portsmouth.

Evaluation at University Hospitals Birmingham NHS Foundation (UHB) trust considered all patients admitted to The Queen Elizabeth Hospital, Birmingham, between December 01, 2019 and October 29, 2020. The Queen Elizabeth Hospital is a large tertiary referral unit within the UHB group which provides healthcare services for a population of 2.2 million across the West Midlands. Confirmatory COVID-19 testing was performed by laboratory SARS-CoV-2 RT-PCR assay.

Evaluation at Bedfordshire NHS Foundation Trust (BHT) considered all patients admitted to Bedford Hospital between January 1, 2021 and March 31, 2021. BHT provides healthcare services for a population of around 620,000 in Bedfordshire. Confirmatory COVID-19 testing was performed on the day of admission by point-of-care PCR based nucleic acid testing [SAMBA-II & Panther Fusion System, Diagnostics in the Real World, UK, and Hologic, USA]. In an evaluation of the SAMBA-II against laboratory RT-PCR testing, the SAMBA-II achieved sensitivity of 96.9% and specificity of 99.1%^9,45^.

We report sensitivity, specificity, positive and negative predictive values (PPV and NPV), AUROC and F1 alongside 95% CIs (Supplementary Table S5 & Figure 2), comparing model predictions to results of confirmatory viral testing (laboratory PCR and SAMBA-II).. Confidence intervals for sensitivity, specificity and predictive values were computed using Wilson’s Method^33^, and for AUROC with DeLong’s method^34^.

**Supplementary Table S5:**
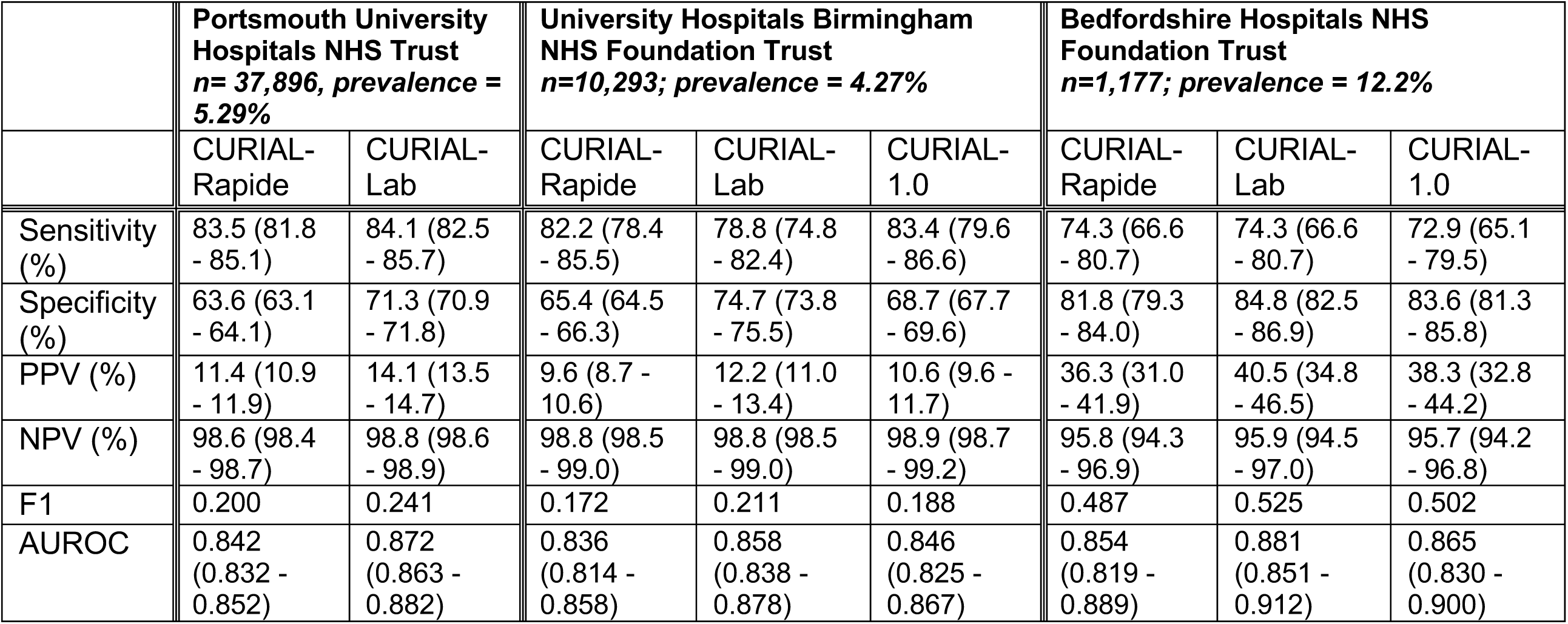
Performance of CURIAL-Rapide, CURIAL-Lab & CURIAL-1.0 (Soltan et al.) during external validation at three UK Hospitals trusts. All models were calibrated during training to achieve 90% sensitivity. Results are reported alongside 95% confidence intervals. (Acronyms – FBC: Complete Blood Count, U&E: Creatinine & Electrolytes, LFD: Liver Function Test, CRP: C-Reactive Protein)

### Comparison with Lateral Flow Tests

We considered any positive lateral flow test which was followed by a positive PCR test within a +/- 48hr window of a patient being admitted to hospital to represent a true positive infection. As previously, model predictions were generated using blood tests performed from the first blood draw on arrival and first-recorded vital signs. In the integrated clinical pathway (Figure 1), patients were considered COVID-19-suspected if they had either a positive LFD result or CURIAL prediction. Results are show in Figure 3 and Supplementary Table S6.

**Supplementary Table S6:**
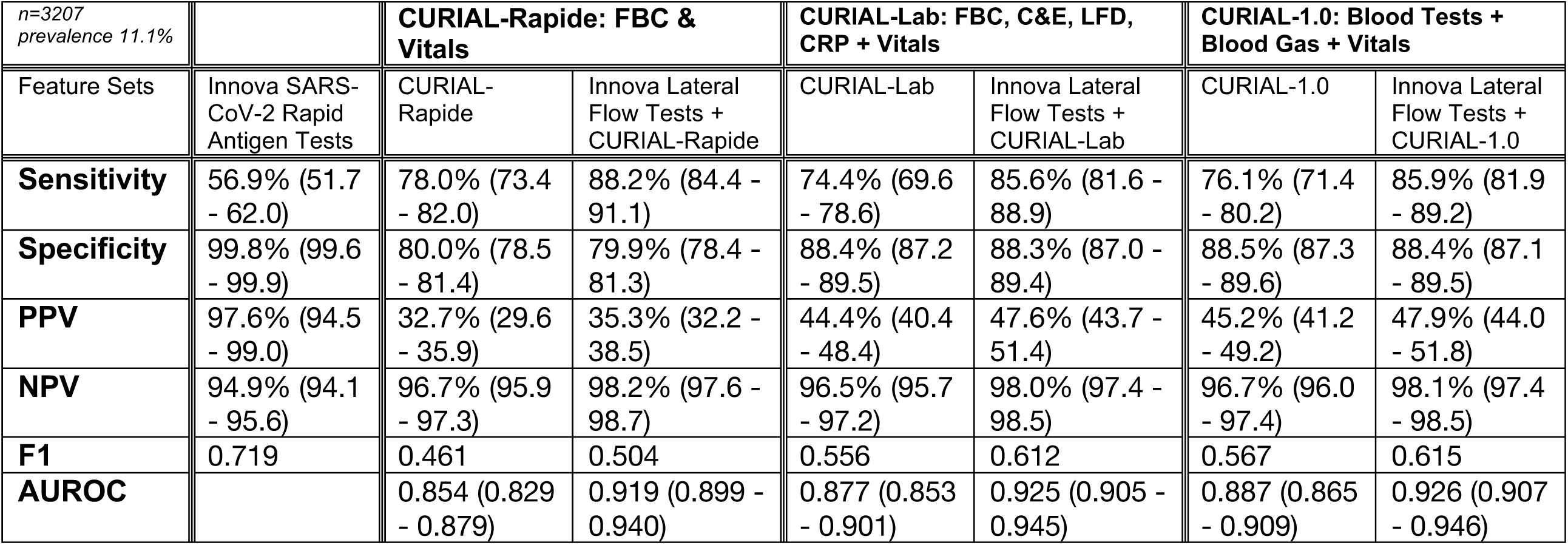
Performance characteristics of (a) INNOVA SARS-CoV2 Rapid Antigen Tests, (b) CURIAL-Rapide & CURIAL-Lab, calibrated during training to a sensitivity of 80%, and (c) combined clinical pathways considering either a positive CURIAL-Rapide/CURIAL-Lab result or a positive LFD test as a COVID-19 suspected case, at Oxford University Hospitals NHS Foundation Trust between December 23, 2020 & March 6, 2021. Error bars show 95% confidence intervals.

## Appendix D: CURIAL-Rapide lab-free service evaluation

Service evaluation of the OLO haematology analyser/CURIAL-Rapide operated between February 18, 2021 and May 10, 2021 between 8am and 8pm.

### Operator Training

We specified that clinical staff carrying out the service evaluation must ordinarily be employed by OUH, participate in the care of patients as part of their usual duties, have completed all statutory & mandatory training required by the trust for their role including for electronic health record systems, and be familiar and competent in using these systems as part of their usual role. We permitted student doctors meeting the above requirements to participate. Training to operate the OLO was provided by in-person device training, supported by demonstration and documentation from the device manufacturers, and a supporting online training video (made available at https://youtu.be/UofBAL7sAzc). Weekly quality-control checks were performed on the OLO analysers.

### Enrolment

OUH sites for eligibility: John Radcliffe Hospital

Inclusion: Adult patients (aged >18)

Clinical areas for sampling eligibility were ED Assessment area, ED Majors Beds and ED Resus. Patients who are not receiving blood tests on presentation to the emergency department as part of their care were not eligible.

### Process

Eligible patients were identified to take part in the service evaluation using the locally-adopted Cerner FirstNet system. Vital signs and blood draws were performed on arrival to the emergency department by healthcare professionals as part of routine care. Following trust procedures, vital signs were documented on the trust electronic health record [SEND; Sensyne Health], and blood bottles were labelled using printed labels from the electronic record. Two drops of venous blood (27uL) from a routinely-collected EDTA blood tube were extracted using a single-use sampling device, and prepared for OLO analysis by trained operators directed by on-screen instructions^31^. OLO results were uploaded immediately to the electronic medical record using the POCcelerator Data Management System [Siemens Healthineers GmbH, Erlangen, Germany], making results available to clinicians and supporting routine patient care. Routine laboratory FBC analysis [Sysmex XN Automated, Sysmex UK] was used to confirm point of care results. Clinical care followed existing pathways and departmental procedures.

**Supplementary Figure S3:**
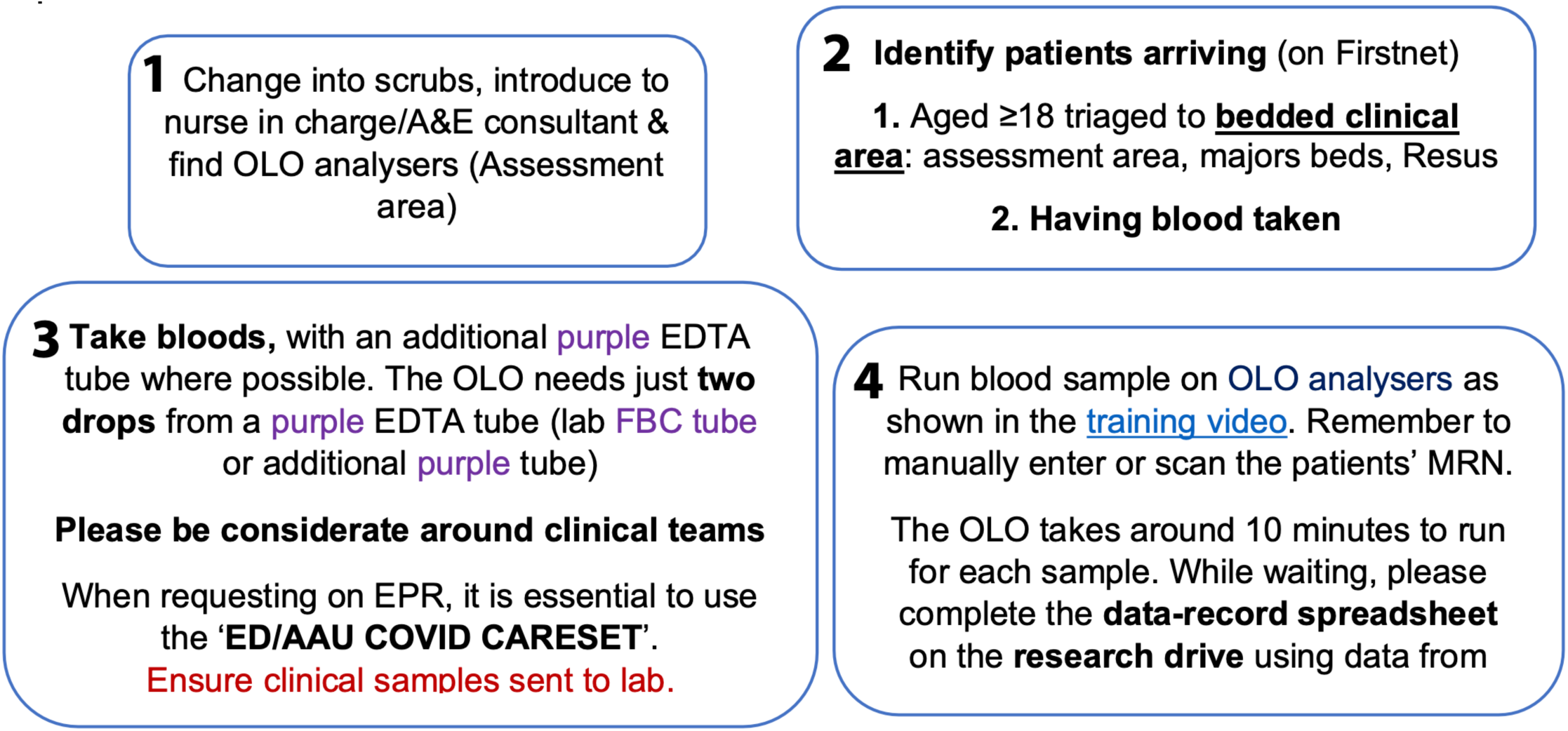
Instructions to trained operators, specifying eligibility criteria for the service evaluation, sample handling and processing techniques.

### Confirmatory COVID-19 Testing

Confirmatory testing of patients enrolled in the OLO/CURIAL-Rapide service evaluation, and LFD comparison, followed OUH trust policies. Swabs of the nose and throat were routinely performed in the emergency department for all patients being admitted to OUH. Lateral Flow Testing (Innova SARS-CoV-2 Antigen Rapid Qualitative Test) was performed in the department, by trained nursing or medical staff, and results were documented on the electronic record. Swabs for PCR were transferred to the clinical laboratory in viral transport medium and tested by PCR (ThermoFisher TaqPath). Where patients were not tested for COVID-19 by confirmatory PCR, or did not receive blood tests or vital signs as part of routine care, we excluded the patients from the CURIAL-Rapide evaluation. We also excluded patients with an invalid OLO result and no subsequent successful result, thereby ensuring data completeness.

### Analysis

Binary CURIAL-Rapide triage predictions (COVID-19-Suspected and COVID-19-Negative) were generated using a custom Python 3.0 application. Libraries used included scikit-learn, pandas, and NumPy. No other clinical data was made available to the algorithm. CURIAL-Rapide predictions were not made available to clinicians in this study, so as not to influence the clinical triage category or decisions to proceed to confirmatory testing.

We compared CURIAL-Rapide predictions, lateral flow results, and clinical triage category by first-assessing clinician against a PCR reference standard. We determined and report sensitivity, specificity, PPV, NPV and accuracy, alongside 95% confidence intervals. We calculated the time-to-result for each test, presenting mean with standard deviation for normally distributed data, and median with interquartile range for data with a skewed distributed (Table 6). Laboratory FBC samples were not processed for 2 of the 520 patients, owing to sample or labelling errors. For paired samples, we compared time-to-result between each test using a one-tailed Wilcoxon Signed Rank test. We additionally performed a Kaplan-Meier survival analysis (Figure 4).

**Table 6:**
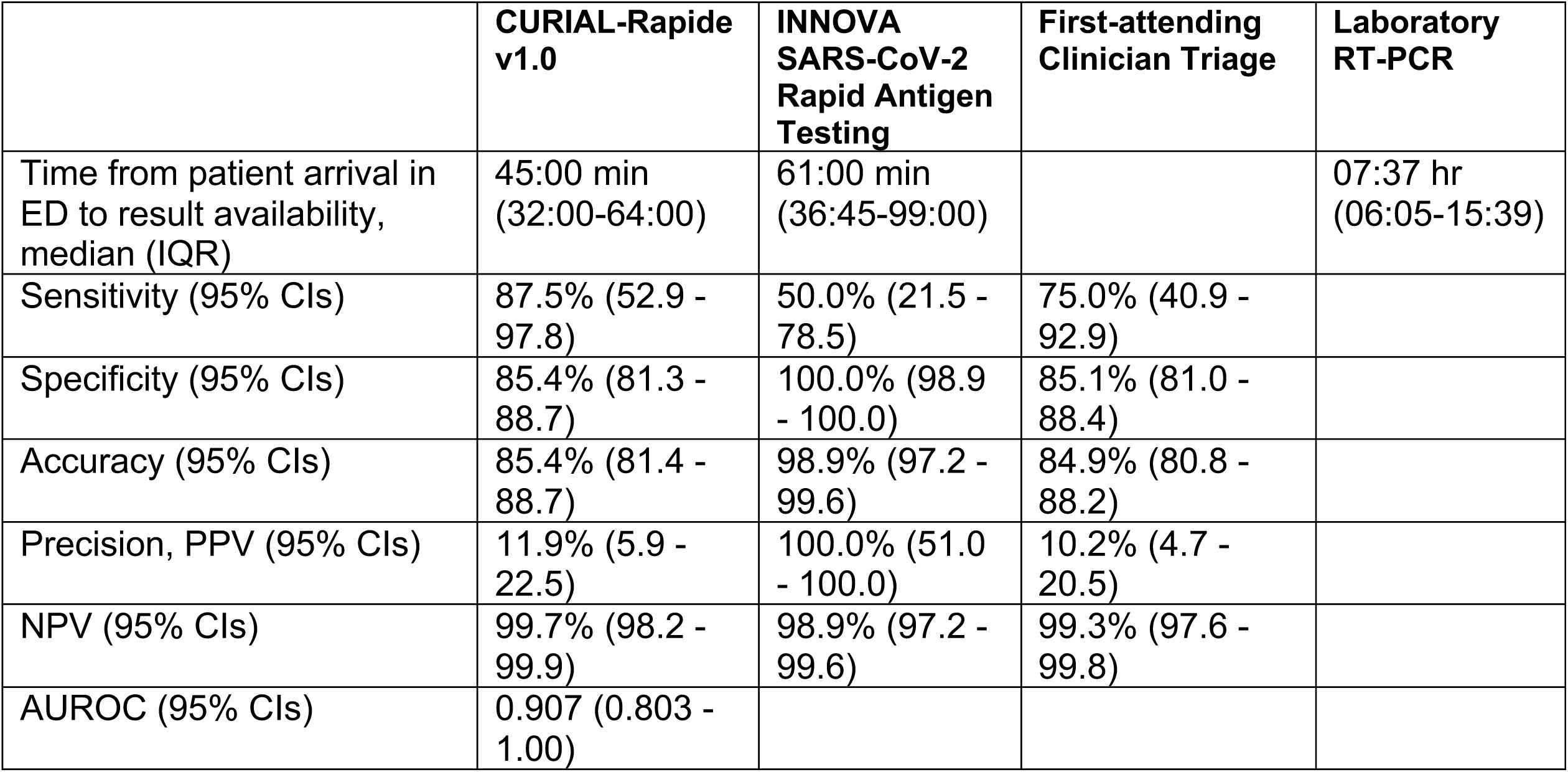
Operational and performance characteristics of (a) CURIAL-Rapide, (b) INNOVA SARS-CoV-2 Rapid Antigen Testing and (c) clinical triage by the first-attending clinician calculated against laboratory RT-PCR testing. Results are reported alongside interquartile range (time-to-result) or 95% confidence intervals.

|  | CURIAL-Rapide v1.0 | INNOVA SARS-CoV-2 Rapid Antigen Testing | First-attending Clinician Triage | Laboratory RT-PCR |
| --- | --- | --- | --- | --- |
| Time from patient arrival in ED to result availability, median (IQR) | 45:00 min (32:00-64:00) | 61:00 min (36:45-99:00) |  | 07:37 hr (06:05-15:39) |
| Sensitivity (95% CIs) | 87.5% (52.9 - 97.8) | 50.0% (21.5 - 78.5) | 75.0% (40.9 - 92.9) |  |
| Specificity (95% CIs) | 85.4% (81.3 - 88.7) | 100.0% (98.9 - 100.0) | 85.1% (81.0 - 88.4) |  |
| Accuracy (95% CIs) | 85.4% (81.4 - 88.7) | 98.9% (97.2 - 99.6) | 84.9% (80.8 - 88.2) |  |
| Precision, PPV (95% CIs) | 11.9% (5.9 - 22.5) | 100.0% (51.0 - 100.0) | 10.2% (4.7 - 20.5) |  |
| NPV (95% CIs) | 99.7% (98.2 - 99.9) | 98.9% (97.2 - 99.6) | 99.3% (97.6 - 99.8) |  |
| AUROC (95% CIs) | 0.907 (0.803 - 1.00) |  |  |  |

We report our study in compliance with the “Standards for Reporting Diagnostic accuracy studies” (STARD) standards^46,47^.

