## Supplementary Information for "Real-world evaluation of AI-driven COVID-19 triage for emergency admissions: External validation & operational assessment of lab-free and high-throughput screening solutions"

Andrew A.S. Soltan et al.

#### Appendix A

#### CURIAL-Rapide Translational Collaborative

We thank all healthcare professionals and students who have supported the CURIAL-Rapide Service evaluation. In particular, we wish to acknowledge:

Adam Watson
Akshay Bhargav
Alex Tough
Alice Rogers
Ayisha M.A. Shaikh
Carolina Valensise
Charlotte Lee
Claire Otasowie
David Metcalfe
Ekta Agarwal
Elham Zareh
Evelyn Thangaraj
Florence Pickles
Gabriella Kelly
Gayatri Tadikamalla
George Shaw
Heather Tong
Hettie Davies
Jaspreet Bahra
Jessica Morgan
Joe Wilson
Joseph Cutteridge
Katherine O'Byrne
Luiza Farache Trajano
Madeleine Oliver
Maria Pikoula
Maya Mendoza
Melissa Keevil
Muhammad Faisal
Natasha Dole
Oscar Deal
Rebecca Conway-Jones
Shajeel Sattar
Sneha Kundoor
Sumaiyah Shah
Vani Muthusami

#### Appendix B:

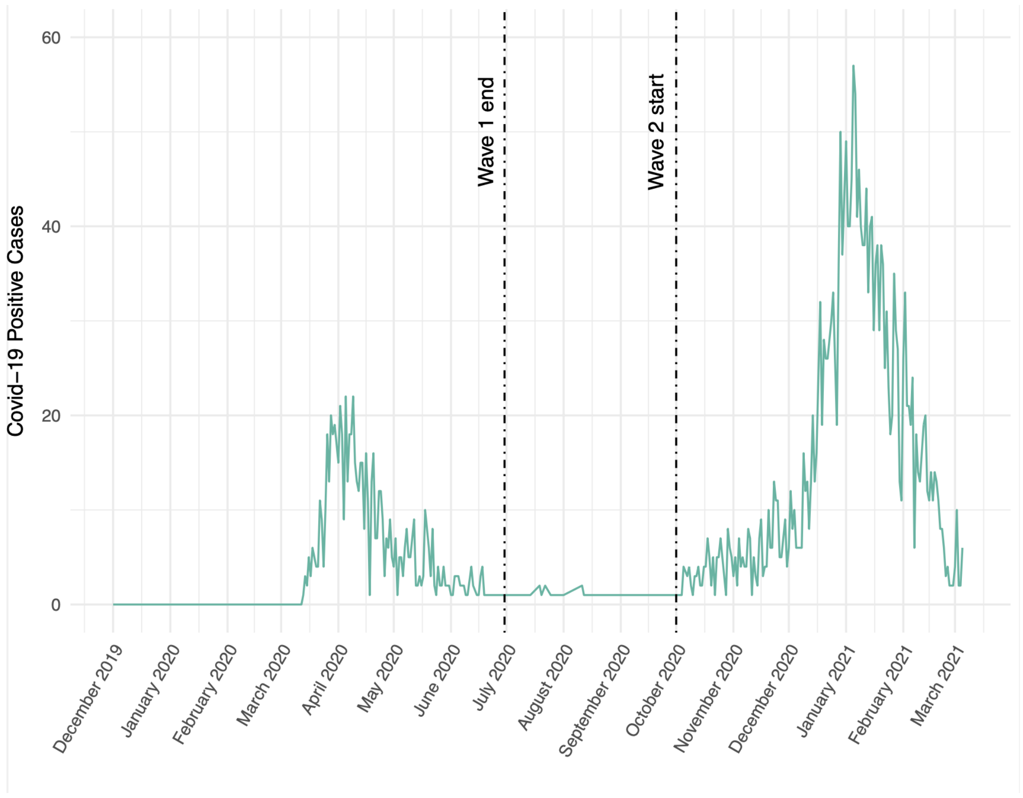


**Supplementary Figure S1:** Daily number of patients presenting to Oxford University Hospitals NHS Foundation Trust testing positive for COVID-19, between 1st December 2019 and 8th March 2021.

**Model Development: Inclusion & Exclusion Criteria**
As previously, we included all patients attending acute and emergency care settings at Oxford University Hospitals NHS foundation trust who received routine blood tests on arrival, considering presentations before December 1, 2019, and thus before the pandemic, as the COVID-19-negative (control) cohort. We considered presentations during the 'first wave' of the UK COVID-19 pandemic (December 1, 2019 to June 30, 2020) with PCR confirmed SARS-CoV-2 infection as the COVID-19-positive (cases) cohort. We excluded patients who opted out of electronic health record (EHR) research and those who did not receive laboratory blood tests or were younger than 18 years of age. Due to incomplete penetrance of testing during the first wave of the pandemic, and imperfect sensitivity of the PCR test, there is uncertainty in the viral status of patients presenting during the pandemic who were untested or tested negative. We therefore selected a pre-pandemic control cohort during training to ensure absence of disease in patients labelled as COVID-19-negative.
Clinical features extracted for each presentation included first-performed blood tests, blood gases, vital signs measurements and PCR testing for SARS-CoV-2 (Abbott Architect [Abbott, Maidenhead, UK], TaqPath [Thermo Fisher Scientific, Massachusetts, USA] and Public Health England-designed RNA-dependent RNA polymerase assays). A list of extracted clinical features is shown in Supplementary Table S1.

**Supplementary Table S1:** Clinical data fields extracted from training and externally validating NHS sites, for all patients admitted to the trusts from 1st December 2019 onwards, and for up to 200,000 pre-pandemic admissions.

| **Clinical Descriptors:** | **Presentation Blood Tests:** | **Presentation Blood Gas:** | **Premorbid Clinical Data** |
|---|---|---|---|
| Study ID | PresentationHAEMOGLOBIN | PresentationPOCT pC02 | BaselineHAEMOGLOBIN |
| Presentation Date | PresentationWHITE CELLS | PresentationPOCT sO2 | BaselineWHITE CELLS |
| Ethnicity | PresentationPLATELETS | PresentationPOCT pO2 | BaselinePLATELETS |
| Age at presentation | PresentationMEAN CELL VOL. | PresentationPCT cBASE(Ecf)c | BaselineMEAN CELL VOL. |
| Gender (M/F) | PresentationRED CELL COUNT | PresentationPCT CO3(P,st)c | BaselineRED CELL COUNT |
| Comorbidities (ICD10) | PresentationNEUTROPHILS | PresentationPOCT Hctc | BaselineNEUTROPHILS |
| Outcome | PresentationHAEMATOCRIT | PresentationPOCT FO2Hb | BaselineHAEMATOCRIT |
| **Vital Signs:** | PresentationLYMPHOCYTES | PresentationPOCT ctO2c | BaselineLYMPHOCYTES |
| AdmissionRespRate | PresentationMEAN CELL HGB | PresentationPOCT cGLU | BaselineMEAN CELL HGB |
| AdmissionHeartRate | PresentationMONOCYTES | PresentationPOCT cK+ | BaselineMONOCYTES |
| AdmissionBloodPressure | PresentationEOSINOPHILS | PresentationPOCT cNA+ | BaselineEOSINOPHILS |
| AdmissionSpO2 | PresentationBASOPHILS | PresentationPOCT cLAC | BaselineBASOPHILS |
| AdmissionOxygenDeliveryDevice | Presentation MCH | PresentationPOCT cCA++ | BaselineMEAN CELL HGB CONC |
| AdmissionTemperature | PresentationMPV | | BaselineSODIUM |
| **Microbiology:** | PresentationNRBC A | | BaselineALBUMIN |
| SARS-CoV-2 PCR | PresentationNRBC % | | BaselineALK.PHOSPHATASE |
| SARS-CoV-2 RESULT TYPE | PresentationSODIUM | | BaselineALT |
| SARS-CoV-2 Antigen Test Result | PresentationALBUMIN | | BaselineUREA |
| INFLUENZAPCR | PresentationALK.PHOSPHATASE | | BaselineBILIRUBIN |
| RespiratoryPCR (Biofire) | PresentationALT | | BaselineCREATININE |
| | PresentationUREA | | BaselineeGFR |
| | PresentationBILIRUBIN | | BaselinePOTASSIUM |
| | PresentationCREATININE | | BaselineCALCIUM |
| | PresentationeGFR | | BaselineADJUSTED CALC. |
| | PresentationPOTASSIUM | | BaselineCRP |
| | PresentationCALCIUM | | BaselineProthromb. Time |
| | PresentationADJUSTED CALC. | | BaselineAPTT |
| | PresentationPHOSPHATE | | BaselineINR |
| | PresentationCRP | | BaselinePOCT pC02 |
| | PresentationProthromb. Time | | BaselinePOCT sO2 |
| | PresentationPOCT ctHb | | BaselinePOCT pO2 |
| | PresentationGLUCOSE | | BaselinePCT cBASE(Ecf)c |
| | PresentationAPTT | | BaselinePCT CO3(P,st)c |
| | PresentationINR | | BaselinePOCT Hctc |
| | | | BaselinePOCT FO2Hb |
| | | | BaselinePOCT ctO2c |
| | | | BaselinePOCT Cglu |
| | | | BaselinePOCT cK+ |
| | | | BaselinePOCT cNA+ |
| | | | BaselinePOCT cLAC |
| | | | BaselinePOCT cCA++ |

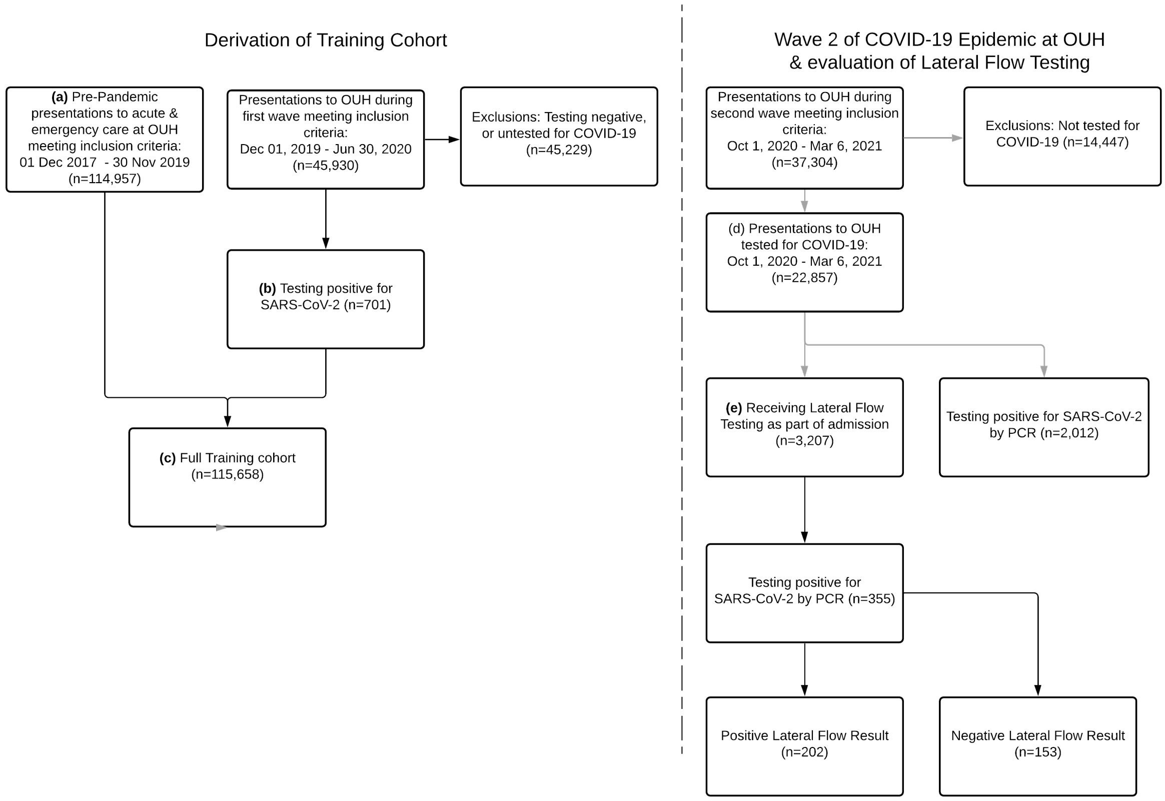


**Supplementary Figure S2:** Participant flow diagram showing patients attending OUH, who met inclusion and exclusion criteria, for (a) the pre-pandemic training cohort and (b) COVID-19-cases cohort, combining to form (c) a full training cohort for model development. Patients attending OUH during the second wave of the UK COVID-19 epidemic, between Oct 1, 2020 and Mar 6, 2020, meeting inclusion and exclusion criteria, formed (d) the second wave analysis cohort, of which a subset (e) received Lateral Flow Testing within routine care, as part of an admission.

**Supplementary Table S2:** Distribution of vital signs, reported as median and interquartile ranges, for each patient cohort.

| | **Training** | | **Prospective Test** | **External Validation (Admissions)** | | | **LFD Evaluation** | **Lab-free Evaluation** |
|---|---|---|---|---|---|---|---|---|
| | Oxford University Hospitals (pre-pandemic & wave 1 cases, to 30 June 2020) | | Oxford University Hospitals | Portsmouth University Hospital NHS Trust | University Hospitals Birmingham NHS Foundation Trust | Bedfordshire Hospitals NHS Foundation Trust | Oxford University Hospitals (wave 2 receiving LFDs) | Oxford University Hospitals ED |
| | Prepandemic cohort | COVID-19-cases cohort | October 1, 2020 – March 6, 2021 | March 1, 2020 - February 28, 2021 | December 01, 2019 - October 29, 2020 | January 1, 2021 - March 31, 2021 | December 23, 2020 – March 6, 2021 | Feb 18, 2021 – May, 10, 2021 |
| **Respiratory Rate (breath/min)** | 18.0 (16.0-19.0) | 20.0 (18.0-24.0) | 18.0 (16.6-19.0) | 17.0 (16.0-19.0) | 18.0 (17.0-20.0) | 18.0 (16.0-20.0) | 18.0 (17.0-20.0) | 18.0 (17.0-20.0) |
| **Heart Rate (beats/min)** | 82.0 (71.0-96.0) | 88.0 (75.0-101.0) | 84.0 (72.0-97.0) | 82.0 (71.0-95.0) | 86.0 (73.0-101.0) | 84.0 (73.0-97.0) | 87.0 (75.0-101.0) | 82.0 (70.0-97.0) |
| **Systolic Blood Pressure (mmHg)** | 132.0 (118.0-150.0) | 131.0 (115.0-146.0) | 134.0 (119.0-152.0) | 128.0 (114.0-146.0) | 136.0 (119.0-155.0) | 131.0 (116.0-149.0) | 136.0 (120.0-156.0) | 146.5 (126.0-168.0) |
| **Diastolic Blood Pressure (mmHg)** | 74.0 (65.0-84.0) | 74.0 (64.0-84.0) | 75.0 (65.0-85.0) | 76.0 (67.0-84.0) | 77.0 (68.0-87.0) | 78.0 (68.0-88.0) | 76.0 (65.0-87.0) | 79.0 (68.0-90.8) |

| | | | | | | | | |
|---|---|---|---|---|---|---|---|---|
| **Oxygen Saturation (%)** | 97.0 (96.0-99.0) | 96.0 (94.0-97.0) | 97.0 (96.0-97.9) | 97.0 (95.0-98.0) | 97.0 (95.0-98.0) | 97.0 (96.0-99.0) | 97.0 (95.0-97.7) | 97.0 (96.0-99.0) |
| **Tympanic Temperature (C)** | 36.5 (36.1-36.9) | 36.9 (36.3-37.6) | 36.3 (36.0-36.7) | 36.3 (36.0-36.8) | 36.7 (36.4-37.2) | 36.5 (36.4-36.9) | 36.3 (35.9-36.8) | 36.3 (35.8-36.8) |

**Supplementary Table S3:** Distribution of blood test features, reported as median and interquartile ranges, for each patient cohort.

| | **Training** | | **Prospective Test** | **External Validation (Admissions)** | | | **LFD Evaluation** | **Lab-free Evaluation** |
|---|---|---|---|---|---|---|---|---|
| | Oxford University Hospitals (pre-pandemic & wave 1 cases, to 30 June 2020) | | Oxford University Hospitals | Portsmouth University Hospital NHS Trust | University Hospitals Birmingham NHS Foundation Trust | Bedfordshire Hospitals NHS Foundation Trust | Oxford University Hospitals (wave 2 receiving LFDs) | Oxford University Hospitals ED (OLO FBC results) |
| | Prepandemic cohort | COVID-19-cases cohort | October 1, 2020 – March 6, 2021 | March 1, 2020 - February 28, 2021 | December 1, 2019 - October 29, 2020 | January 1, 2021 - March 31, 2021 | December 23, 2020 – March 6, 2021 | Feb 18, 2021 – May, 10, 2021 |
| **HAEMOGLOBIN (g/L)** | 130.0 (116.0-142.0) | 130.0 (114.0-144.0) | 129.0 (114.0-142.0) | 129.0 (114.0-143.0) | 127.0 (113.0-140.0) | 134.0 (119.0-146.0) | 131.0 (116.0-144.0) | 125.0 (112.0-137.5) |
| **WHITE CELLS ($10^9$ $l^{-1}$)** | 8.45 (6.46-11.18) | 6.98 (5.14-9.72) | 8.94 (6.7-12.06) | 8.6 (6.7-11.3) | 9.4 (7.1-12.6) | 9.2 (6.9-12.5) | 9.43 (7.05-12.73) | 8.56 (6.68-11.37) |
| **PLATELETS ($10^9$ $l^{-1}$)** | 249.0 (199.0-307.0) | 215.0 (163.0-283.5) | 251.0 (198.0-314.0) | 251.0 (199.0-312.0) | 247.0 (196.0-311.0) | 246.0 (196.0-310.0) | 249.0 (195.0-313.0) | 223.0 (183.5-270.5) |
| **MEAN CELL VOL (fl)** | 89.6 (86.0-93.4) | 90.2 (86.6-94.2) | 90.2 (86.6-94.2) | 89.0 (84.9-93.0) | 89.9 (86.2-93.6) | 88.0 (85.0-92.0) | 90.0 (86.4-94.3) | 90.2 (87.1-93.7) |
| **NEUTROPHILS ($10^9$ $l^{-1}$)** | 5.72 (3.99-8.36) | 5.11 (3.48-7.49) | 6.44 (4.4-9.55) | 5.9 (4.2-8.6) | 6.9 (4.7-10.0) | 6.8 (4.7-9.73) | 6.97 (4.68-10.19) | 6.26 (4.36-9.05) |
| **HAEMATOCRIT** | 0.39 (0.35-0.42) | 0.4 (0.35-0.44) | 0.39 (0.35-0.43) | 0.39 (0.34-0.42) | 0.38 (0.34-0.42) | 0.39 (0.35-0.43) | 0.4 (0.36-0.43) | 0.37 (0.33-0.41) |
| **LYMPHOCYTES ($10^9$ $l^{-1}$)** | 1.51 (1.0-2.13) | 0.96 (0.65-1.38) | 1.31 (0.85-1.89) | 1.5 (0.97-2.2) | 1.3 (0.9-1.9) | 1.27 (0.86-1.83) | 1.26 (0.83-1.89) | 1.25 (0.86-1.78) |
| **MONOCYTES ($10^9$ $l^{-1}$)** | 0.64 (0.48-0.85) | 0.49 (0.35-0.74) | 0.66 (0.48-0.89) | 0.63 (0.48-0.85) | 0.7 (0.5-0.9) | 0.66 (0.48-0.92) | 0.68 (0.49-0.93) | 0.59 (0.43-0.78) |
| **EOSINOPHILS ($10^9$ $l^{-1}$)** | 0.1 (0.04-0.2) | 0.01 (0.0-0.06) | 0.07 (0.02-0.16) | 0.1 (0.02-0.2) | 0.1 (0.0-0.2) | 0.06 (0.02-0.16) | 0.06 (0.01-0.14) | 0.09 (0.05-0.17) |
| **BASOPHILS ($10^9$ $l^{-1}$)** | 0.04 (0.03-0.06) | 0.02 (0.01-0.03) | 0.04 (0.02-0.06) | 0.04 (0.02-0.06) | 0.1 (0.0-0.1) | 0.05 (0.03-0.07) | 0.04 (0.02-0.06) | 0.03 (0.01-0.04) |
| **SODIUM (mM)** | 138.0 (136.0-140.0) | 136.0 (134.0-139.0) | 138.0 (135.0-140.0) | 138.0 (136.0-140.0) | 137.0 (134.0-139.0) | 138.0 (136.0-140.0) | 138.0 (135.0-140.0) | |
| **ALBUMIN (g/L)** | 36.0 (32.0-39.0) | 32.0 (28.0-35.0) | 36.0 (31.0-39.0) | 36.0 (31.0-40.0) | 36.0 (32.0-40.0) | 35.0 (31.0-39.0) | 36.0 (31.0-39.0) | |
| **ALKALINE PHOSPHATASE (IU/L)** | 80.0 (64.0-105.0) | 82.0 (64.0-108.0) | 84.0 (66.0-112.0) | 84.0 (67.0-109.0) | 90.0 (71.0-119.0) | 94.0 (74.5-122.0) | 86.0 (69.0-115.0) | |
| **ALT (IU/L)** | 18.0 (13.0-28.0) | 25.0 (17.0-41.0) | 20.0 (13.0-33.0) | 19.0 (13.0-30.0) | 19.0 (13.0-30.0) | 20.0 (13.0-31.0) | 20.0 (13.0-33.0) | |
| **UREA (mM)** | 5.3 (4.0-7.4) | 5.9 (4.2-9.07) | 5.7 (4.2-8.3) | 5.2 (3.8-7.6) | 6.2 (4.5-9.0) | 5.8 (4.2-8.3) | 5.9 (4.3-8.8) | |
| **BILIRUBIN (umol/L)** | 9.0 (6.0-13.0) | 9.0 (7.0-13.25) | 9.0 (6.0-14.0) | 10.0 (7.0-16.0) | 10.0 (7.0-15.0) | 10.0 (7.0-14.0) | 10.0 (7.0-14.0) | |
| **CREATININE (umol/L)** | 73.0 (60.0-93.0) | 79.0 (65.0-106.0) | 74.0 (60.0-97.0) | 74.0 (60.0-96.0) | 78.0 (62.0-105.0) | 80.5 (65.75-104.0) | 74.0 (60.0-98.0) | |
| **eGFR (ml/min)** | 85.0 (63.0-150.0) | 78.0 (53.0-150.0) | 84.0 (58.0-150.0) | 83.0 (60.0-90.0) | 76.0 (52.0-90.0) | 76.0 (54.0-90.0) | 82.0 (56.0-150.0) | |
| **POTASSIUM (mM)** | 4.0 (3.7-4.3) | 4.0 (3.7-4.3) | 4.0 (3.8-4.4) | 4.2 (3.9-4.4) | 4.1 (3.8-4.4) | 4.3 (4.0-4.6) | 4.1 (3.8-4.4) | |
| **CRP (mg/L)** | 8.6 (2.3-39.0) | 72.5 (23.8-143.6) | 15.8 (3.5-67.4) | 13.0 (3.0-71.0) | 12.0 (3.0-61.0) | 10.7 (2.8-48.78) | 17.9 (3.6-77.5) | |

**Supplementary Table S4:** Distribution of blood gas features, reported as median and interquartile ranges for each patient cohort.

| | **Training** | | **Prospective Test** | **External Validation (Admissions)** | | **LFD Evaluation** |
|---|---|---|---|---|---|---|
| | Oxford University Hospitals (pre-pandemic & wave 1 cases, to 30 June 2020) | | Oxford University Hospitals | University Hospitals Birmingham NHS Foundation Trust | Bedfordshire Hospitals NHS Foundation Trust | Oxford University Hospitals (wave 2 receiving LFDs) |
| | Prepandemic cohort | COVID-19-cases cohort | October 1, 2020 – March 6, 2021 | December 01, 2019 - October 29, 2020 | January 1, 2021 - March 31, 2021 | December 23, 2020 – March 6, 2021 |
| **pCO2 (kPa)** | 5.57 (4.94-6.22) | 5.34 (4.57-6.01) | 5.61 (4.95-6.28) | 5.7 (5.0-6.5) | 5.72 (5.03-6.43) | 5.68 (5.0-6.4) |
| **O2 Sat (%)** | 64.5 (44.0-83.8) | 65.15 (38.85-84.68) | 65.3 (43.6-85.8) | 69.8 (44.6-89.9) | 68.0 (44.0-88.9) | 60.35 (40.05-80.2) |
| **pO2 (kPa)** | 4.68 (3.51-6.53) | 4.62 (3.4-6.7) | 4.79 (3.54-6.92) | 5.0 (3.4-7.4) | 4.86 (3.41-7.2) | 4.52 (3.44-6.2) |
| **BE Std (mM)** | 1.3 (-0.7-3.2) | 1.3 (-0.8-3.3) | 1.4 (-0.8-3.5) | -0.1 (-2.1-1.5) | 2.2 (0.02-4.2) | 1.6 (-0.6-3.8) |
| **Bicarbonate (mM)** | 24.7 (23.2-26.0) | 24.7 (23.2-26.3) | 24.8 (23.2-26.3) | 24.9 (22.6-27.1) | 27.2 (24.72-29.6) | 24.8 (23.1-26.3) |
| **Haematocrit** | 41.6 (37.2-45.4) | 41.3 (36.8-45.8) | 41.3 (36.4-45.5) | 43.0 (38.7-46.5) | 41.8 (36.9-45.9) | 42.0 (37.3-46.0) |
| **Glucose (mM)** | 6.2 (5.4-7.5) | 6.6 (5.7-8.2) | 6.4 (5.5-8.0) | 6.73 (5.81-8.48) | 6.4 (5.5-8.0) | 6.6 (5.6-8.3) |
| **K+ (mM)** | 3.9 (3.7-4.3) | 3.85 (3.6-4.2) | 4.0 (3.7-4.3) | 3.96 (3.66-4.3) | 4.0 (3.7-4.3) | 4.0 (3.7-4.3) |
| **Na+ (mM)** | 138.0 (135.0-141.0) | 137.0 (133.0-140.0) | 138.0 (135.0-141.0) | 140.0 (137.2-141.9) | 139.0 (135.0-141.0) | 138.0 (135.0-141.0) |
| **cLAC (mM)** | 1.3 (0.9-1.9) | 1.4 (1.24-2.0) | 1.4 (1.24-1.9) | 1.64 (1.25-2.27) | 1.3 (1.0-1.9) | 1.4 (1.24-2.1) |
| **Ca2+ (mM)** | 1.18 (1.14-1.21) | 1.12 (1.08-1.16) | 1.17 (1.13-1.21) | 1.21 (1.16-1.24) | 1.17 (1.12-1.2) | 1.17 (1.14-1.21) |
| **Haemoglobin (g/dL)** | 136.0 (121.0-148.0) | 134.0 (120.0-149.0) | 135.0 (118.0-148.0) | 133.1 (118.4-146.3) | 136.0 (120.0-150.0) | 137.0 (121.0-150.0) |

**Supplementary Table S5:** Performance of CURIAL-Rapide, CURIAL-Lab & CURIAL-1.0 (Soltan et al.) during external validation at three UK Hospitals trusts. All models were calibrated during training to achieve 90% sensitivity. Results are reported alongside 95% confidence intervals. (Acronyms – FBC: Complete Blood Count, U&E: Creatinine & Electrolytes, LFD: Liver Function Test, CRP: C-Reactive Protein)

| | **Portsmouth University Hospitals NHS Trust** ***n= 37,896, prevalence = 5.29%*** | | **University Hospitals Birmingham NHS Foundation Trust** ***n=10,293; prevalence = 4.27%*** | | | **Bedfordshire Hospitals NHS Foundation Trust** ***n=1,177; prevalence = 12.2%*** | | |
|---|---|---|---|---|---|---|---|---|
| | CURIAL-Rapide | CURIAL-Lab | CURIAL-Rapide | CURIAL-Lab | CURIAL-1.0 | CURIAL-Rapide | CURIAL-Lab | CURIAL-1.0 |
| Sensitivity (%) | 83.5 (81.8 - 85.1) | 84.1 (82.5 - 85.7) | 82.2 (78.4 - 85.5) | 78.8 (74.8 - 82.4) | 83.4 (79.6 - 86.6) | 74.3 (66.6 - 80.7) | 74.3 (66.6 - 80.7) | 72.9 (65.1 - 79.5) |
| Specificity (%) | 63.6 (63.1 - 64.1) | 71.3 (70.9 - 71.8) | 65.4 (64.5 - 66.3) | 74.7 (73.8 - 75.5) | 68.7 (67.7 - 69.6) | 81.8 (79.3 - 84.0) | 84.8 (82.5 - 86.9) | 83.6 (81.3 - 85.8) |
| PPV (%) | 11.4 (10.9 - 11.9) | 14.1 (13.5 - 14.7) | 9.6 (8.7 - 10.6) | 12.2 (11.0 - 13.4) | 10.6 (9.6 - 11.7) | 36.3 (31.0 - 41.9) | 40.5 (34.8 - 46.5) | 38.3 (32.8 - 44.2) |
| NPV (%) | 98.6 (98.4 - 98.7) | 98.8 (98.6 - 98.9) | 98.8 (98.5 - 99.0) | 98.8 (98.5 - 99.0) | 98.9 (98.7 - 99.2) | 95.8 (94.3 - 96.9) | 95.9 (94.5 - 97.0) | 95.7 (94.2 - 96.8) |
| F1 | 0.200 | 0.241 | 0.172 | 0.211 | 0.188 | 0.487 | 0.525 | 0.502 |
| AUROC | 0.842 (0.832 - 0.852) | 0.872 (0.863 - 0.882) | 0.836 (0.814 - 0.858) | 0.858 (0.838 - 0.878) | 0.846 (0.825 - 0.867) | 0.854 (0.819 - 0.889) | 0.881 (0.851 - 0.912) | 0.865 (0.830 - 0.900) |

### Comparison with Lateral Flow Tests

We considered any positive lateral flow test which was followed by a positive PCR test within a +/- 48hr window of a patient being admitted to hospital to represent a true positive infection. As previously, model predictions were generated using blood tests performed from the first blood draw on arrival and first-recorded vital signs. In the integrated clinical pathway (Figure 1), patients were considered COVID-19-suspected if they had either a positive LFD result or CURIAL prediction. Results are show in Figure 3 and Supplementary Table S6.

**Supplementary Table S6:** Performance characteristics of (a) INNOVA SARS-CoV2 Rapid Antigen Tests, (b) CURIAL-Rapide & CURIAL-Lab, calibrated during training to a sensitivity of 80%, and (c) combined clinical pathways considering either a positive CURIAL-Rapide/CURIAL-Lab result or a positive LFD test as a COVID-19 suspected case, at Oxford University Hospitals NHS Foundation Trust between December 23, 2020 & March 6, 2021. Error bars show 95% confidence intervals.

| *n=3207 prevalence 11.1%* | | **CURIAL-Rapide: FBC & Vitals** | | **CURIAL-Lab: FBC, C&E, LFD, CRP + Vitals** | | **CURIAL-1.0: Blood Tests + Blood Gas + Vitals** | |
|---|---|---|---|---|---|---|---|
| Feature Sets | Innova SARS-CoV-2 Rapid Antigen Tests | CURIAL-Rapide | Innova Lateral Flow Tests + CURIAL-Rapide | CURIAL-Lab | Innova Lateral Flow Tests + CURIAL-Lab | CURIAL-1.0 | Innova Lateral Flow Tests + CURIAL-1.0 |
| **Sensitivity** | 56.9% (51.7 - 62.0) | 78.0% (73.4 - 82.0) | 88.2% (84.4 - 91.1) | 74.4% (69.6 - 78.6) | 85.6% (81.6 - 88.9) | 76.1% (71.4 - 80.2) | 85.9% (81.9 - 89.2) |
| **Specificity** | 99.8% (99.6 - 99.9) | 80.0% (78.5 - 81.4) | 79.9% (78.4 - 81.3) | 88.4% (87.2 - 89.5) | 88.3% (87.0 - 89.4) | 88.5% (87.3 - 89.6) | 88.4% (87.1 - 89.5) |
| **PPV** | 97.6% (94.5 - 99.0) | 32.7% (29.6 - 35.9) | 35.3% (32.2 - 38.5) | 44.4% (40.4 - 48.4) | 47.6% (43.7 - 51.4) | 45.2% (41.2 - 49.2) | 47.9% (44.0 - 51.8) |
| **NPV** | 94.9% (94.1 - 95.6) | 96.7% (95.9 - 97.3) | 98.2% (97.6 - 98.7) | 96.5% (95.7 - 97.2) | 98.0% (97.4 - 98.5) | 96.7% (96.0 - 97.4) | 98.1% (97.4 - 98.5) |
| **F1** | 0.719 | 0.461 | 0.504 | 0.556 | 0.612 | 0.567 | 0.615 |
| **AUROC** | | 0.854 (0.829 - 0.879) | 0.919 (0.899 - 0.940) | 0.877 (0.853 - 0.901) | 0.925 (0.905 - 0.945) | 0.887 (0.865 - 0.909) | 0.926 (0.907 - 0.946) |

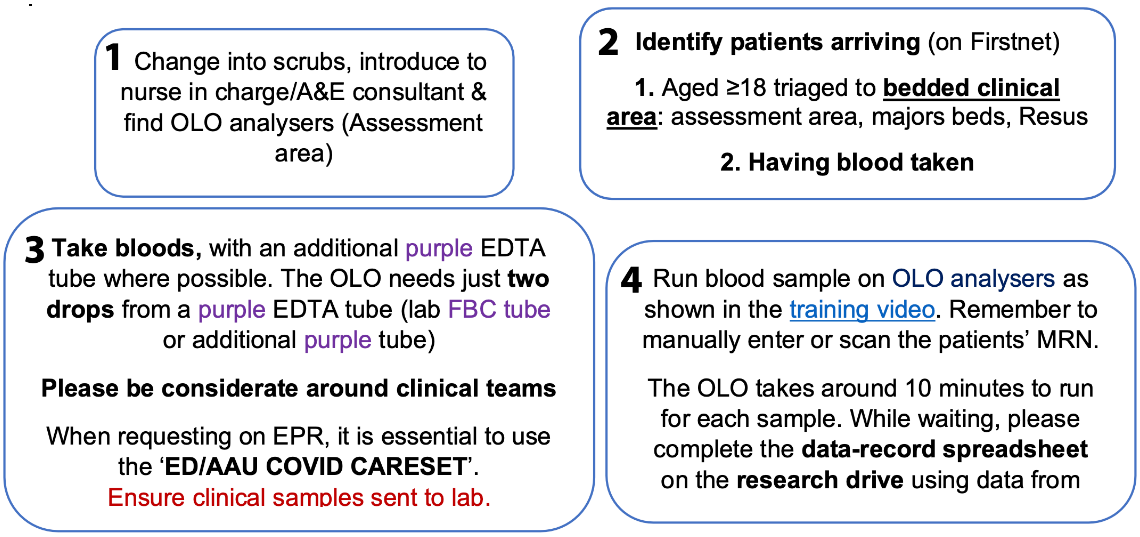


**Supplementary Figure S3:** Instructions to trained operators, specifying eligibility criteria for the service evaluation, sample handling and processing techniques.

### Confirmatory COVID-19 Testing:

Confirmatory testing of patients enrolled in the OLO/CURIAL-Rapide service evaluation, and LFD comparison, followed OUH trust policies. Swabs of the nose and throat were routinely performed in the emergency department for all patients being admitted to OUH. Lateral Flow Testing (Innova SARS-CoV-2 Antigen Rapid Qualitative Test) was performed in the department, by trained nursing or medical staff, and results were documented on the electronic record. Swabs for PCR were transferred to the clinical laboratory in viral transport medium and tested by PCR (ThermoFisher TaqPath). Where patients were not tested for COVID-19 by confirmatory PCR, or did not receive blood tests or vital signs as part of routine care, we excluded the patients from the CURIAL-Rapide evaluation. We also excluded patients with an invalid OLO result and no subsequent successful result, thereby ensuring data completeness.

### Analysis

Binary CURIAL-Rapide triage predictions (COVID-19-Suspected and COVID-19-Negative) were generated using a custom Python 3.0 application. Libraries used included scikit-learn, pandas, and NumPy. No other clinical data was made available to the algorithm. CURIAL-Rapide predictions were not made available to clinicians in this study, so as not to influence the clinical triage category or decisions to proceed to confirmatory testing.

We compared CURIAL-Rapide predictions, lateral flow results, and clinical triage category by first-assessing clinician against a PCR reference standard. We determined and report sensitivity, specificity, PPV, NPV and accuracy, alongside 95% confidence intervals. We calculated the time-to-result for each test, presenting mean with standard deviation for normally distributed data, and median with interquartile range for data with a skewed distributed (Table 6). Laboratory FBC samples were not processed for 2 of the 520 patients, owing to sample or labelling errors. For paired samples, we compared time-to-result between each test using a one-tailed Wilcoxon Signed Rank test. We additionally performed a Kaplan-Meier survival analysis (Figure 4).

We report our study in compliance with the "Standards for Reporting Diagnostic accuracy studies" (STARD) standards[46,47].
